# Cardiovascular risk of dementia is associated with brain–behaviour changes in cognitively healthy, middle-aged individuals

**DOI:** 10.1101/2022.09.02.22279541

**Authors:** Feng Deng, Maria-Eleni Dounavi, Emanuele R. G. Plini, Karen Ritchie, Graciela Muniz-Terrera, Siobhan Hutchinson, Paresh Malhotra, Craig W Ritchie, Brian Lawlor, Lorina Naci

**Affiliations:** Trinity College Institute of Neuroscience, School of Psychology, Trinity College Dublin, Dublin, Ireland; Global Brain Health Institute, Trinity College Dublin, Dublin, Ireland; Department of Psychiatry, School of Clinical Medicine, University of Cambridge, Cambridge CB2 0SP, UK; U1061 Neuropsychiatry, INSERM, University of Montpellier, Montpellier, France; Edinburgh Dementia Prevention, University of Edinburgh, Edinburgh, UK; Department of Social medicine, Ohio University, USA; Department of Neurology, St James’s Hospital, Dublin, Ireland; Department of Brain Science, Imperial College Healthcare NHS Trust, UK

**Keywords:** Alzheimer’s disease, Cognition, Locus Coeruleus, Hippocampus, Functional connectivity, Midlife, Cardiovascular risk score

## Abstract

Alzheimer’s Disease (AD) neuropathology start decades before clinical manifestations, but whether risk factors are associated with early cognitive and brain changes in midlife remains poorly understood. We examined whether AD risk factors were associated with cognition and functional connectivity (FC) between the Locus Coeruleus (LC) and hippocampus – two key brain structures in AD neuropathology – cross-sectionally and longitudinally in cognitively healthy midlife individuals. Neuropsychological assessments and functional Magnetic Resonance Imaging were obtained at baseline (N=210), and two-years follow-up (N=188). Associations of cognition and FC with apolipoprotein ε4 (APOE ε4) genotype, family history of dementia, and the Cardiovascular Risk Factors, Aging, and Incidence of Dementia (CAIDE) score were investigated. Cross-sectionally, higher CAIDE scores were associated with worse cognition. Menopausal status interacted with the CAIDE risk on cognition. Furthermore, the CAIDE score significantly moderated the relationship between cognition and LC–Hippocampus FC. Longitudinally, the LC–Hippocampus FC decreased significantly over 2 years. These results shed light on some of the earliest effects of AD risk on cognition and the brain.

## 1. Introduction

Dementia, particularly Alzheimer’s disease (AD), is a growing epidemic that presents profound challenges to healthcare systems, families, and societies throughout the world (World Health Organization, 2021). Midlife is a critical period for the beginning of AD pathology (Jansen et al., 2015; Sperling et al., 2011) and potentially a unique disease-altering window prior to the manifestation of substantial brain damage. Therefore, there is an urgent need for risk reduction interventions focused on midlife (Barnes & Yaffe, 2011; Livingston et al., 2017; Ritchie et al., 2010). However, the indicators and brain mechanism of AD in midlife remain poorly understood (Irwin et al., 2018; Ritchie et al., 2017).

To investigate early changes, studies in preclinical AD populations have used risk stratification approaches. Key risk factors include the Apolipoprotein E (APOE) ε4 genotype (Donix et al., 2012; Liu et al., 2013), the main genetic risk factor for sporadic late-onset AD in the Indo-European population (Lambert et al., 2013), and family history of dementia (FHD) (Berti et al., 2011; Donix et al., 2012; Scarabino et al., 2016). Additionally, there is growing evidence to suggest that up to 40% of all dementia cases are associated with known modifiable risk factors (Livingston et al., 2020). Among dementia risk scores that incorporate lifestyle risk factors (Barnes et al., 2014; Deckers et al., 2015; Kivipelto et al., 2006), the Cardiovascular Risk Factors Aging and Dementia (CAIDE) score has been optimized for midlife (Kivipelto et al., 2006) and validated in a large US population followed longitudinally over 40 years (Exalto et al., 2014).

AD is neuropathologically characterized by the accumulation of beta-amyloid (A*β*) and hyperphosphorylated tau (pTau) (Braak & Braak, 1991; Braak et al., 2011). Recent studies suggest that tau deposition is a key etiological factor that presages sporadic AD (Arnsten et al., 2021; Jacobs et al., 2021; Kametani & Hasegawa, 2018). Analyses of thousands of human brains across the lifespan suggest that tau pathology may begin about a decade before formation of A*β* plaques (Braak & Del Tredici, 2015). Additionally, tau pathology correlates with progressive grey matter loss to a greater extent than A*β* (La Joie et al., 2019) and cognitive impairment (Giannakopoulos et al., 2003). Neuropathological findings show that the nucleus locus coeruleus (LC) is one of the subcortical regions where tau pathology starts in late young adulthood and early midlife (30–40 years) (Gilvesy et al., 2022; Ehrenberg et al., 2017; Braak & Braak, 1991) then spreads to the transentorhinal and entorhinal cortex (Braak et al., 2011), to the hippocampal and neocortical association cortex, and finally throughout the neocortex.

It has been suggested that early tau pathology in the LC induces LC hyperactivity, thereby promoting its own spread to interconnected brain regions and facilitating the progression of the disease (Weinshenker, 2018). A recent review of the neuropathological literature across the lifespan (Arnsten et al., 2021) suggests that the spread of tau pathology from the LC to the medial temporal lobe (MTL) captures the earliest stages of incipient AD progression (but see Kaufman et al. (2018) for a differing account). However, alterations of connectivity between the LC and the hippocampus, due to AD risk factors have not previously been studied in midlife preclinical populations.

LC is the earliest site of tau pathology accumulation, and, furthermore, the main brain site of noradrenaline production (Amaral & Sinnamon 1977), and thus the originating node of the brain’s arousal system (Sara, 2009; Aston-Jones & Cohen 2005). Therefore, early LC pathology can critically impact arousal and cognitive processes throughout the brain (Mather & Harley, 2016; Betts, Kirilina, et al., 2019). Previous in vivo studies of the LC have investigated healthy lifespan adult cohorts (Betts et al., 2017; Clewett et al., 2016; Lee et al., 2020; Liu et al., 2019; Porat et al., 2022), cognitively unimpaired older adults and preclinical populations with risk for sporadic AD (Jacobs et al., 2021; Prokopiou et al., 2022; Van Egroo et al., 2021), individuals with preclinical autosomal dominant Alzheimer’s disease (Jacobs et al., 2022), mild cognitive impairment (Jacobs et al., 2015; Takahashi et al., 2015), and Alzheimer’s disease (Betts, Cardenas-Blanco, et al., 2019; Dahl et al., 2022; Olivieri et al., 2019; Takahashi et al., 2015) and found significant negative associations of LC integrity with age, age-related cognitive decline (Calarco et al., 2022; Dahl et al., 2019; Liu et al., 2020), AD pathology, such as amyloid (Betts, Cardenas-Blanco, et al., 2019) and tau deposition (Dahl et al., 2022; Jacobs et al., 2022), as well as with symptom severity in AD (Sun et al., 2020). Prokopiu et al. (2022) also found that lower novelty-related LC activity and LC to hippocampus and parahippocampus functional connectivity were associated with steeper beta-amyloid-related cognitive decline. However, by contrast to its key importance to cognitive function and etiological role in AD progression, there is very limited in vivo understanding of LC function disruptions during the early preclinical period. One exception is a recent study of asymptomatic midlife adults, offspring of late-onset AD (O-LOAD) patients. Del Cerro and colleagues (2020) found that the connectivity of the LC and cerebellum was significantly associated with delayed recall performance, and reduced in O-LOAD individuals relative to those with no family history.

The hippocampus is one of the first regions to demonstrate atrophy during prodromal AD. Neurofibrillary tangle (NFT) accumulation within the MTL, including the hippocampus, is one of the first hallmark pathological manifestations. Recent studies in cognitively healthy individuals at risk for late-life AD have found that AD risk impacts the hippocampus in midlife (Dounavi et al., 2022; 2020; Kerchner et al., 2014; for a review see Vilor-Tejedor et al. 2021). Dounavi et al. (2020) reported that the volume of the molecular layer of the hippocampus was reduced in cognitively healthy midlife individuals with APOE ε4 genotype. A more recent study (Dounavi et al., 2022) reported that the hippocampal fissure was enlarged in middle-aged individuals with high CAIDE dementia risk scores.

Critically, the connectivity between the LC and the hippocampus is important for cognition. Animal studies have demonstrated that the co-release of noradrenaline and dopamine from LC terminals in the hippocampus is critical for successful spatial learning and memory (James et al., 2021; Kaufman et al., 2020; Kempadoo et al., 2016). Human studies in healthy older adults are consistent with these data and show that higher LC–Hippocampus functional connectivity is significantly associated with better memory (Jacobs et al., 2015). While accumulating evidence indicates that, individually, the LC and hippocampus are affected in midlife by risk of late-life AD, it remains unknown whether their interactions and joint role in cognition are affected by AD risk at this stage. Understanding of changes in the interaction of LC and the hippocampus during this preclinical period in at-risk populations has the potential to not only shed light on the brain mechanisms of incipient AD, but also to provide urgently needed early disease biomarkers.

To address this gap, we investigated the impact of three risk factors for late-life AD (APOE ε4 genotype, FHD and CAIDE score) on cognition, and its relationship to the LC– Hippocampus functional connectivity. We tested a cohort of cognitively healthy middle-aged individuals, at baseline (N=210), and two-years follow-up (N=188) with a detailed neuropsychological battery not restricted to functions usually implicated in dementia detection in the older adults. Structural and functional Magnetic Resonance Imaging (MRI) data was obtained at both research visits. Our hypotheses were that risk of late-life AD would be significantly associated with cognition, the LC–Hippocampus functional connectivity, and their brain–behaviour relationship in middle-aged and cognitively healthy individuals. Furthermore, we expected that risk factors would be associated with longitudinal change of cognition and functional connectivity in this cohort.

## 2. Methods

### 2.1 Participants

PREVENT-Dementia is an ongoing longitudinal multi-site research programme across the UK and Ireland, seeking to identify early biomarkers of AD and elaborate on risk-mechanism interactions for neurodegenerative diseases decades before the cardinal symptoms of dementia emerge. Its protocol has been described in detail elsewhere (Ritchie & Ritchie, 2012). In the first PREVENT programme phase, participants were recruited at a single site, via the dementia register database held at the West London National Health Service (NHS) Trust, of the UK National Health Service, the Join Dementia Research website (https://www.joindementiaresearch.nihr.ac.uk/), through public presentations, social media and word of mouth. Procedures involving experiments on human participants were carried out in accord with the ethical standards of the Institutional Review Board of Imperial College London and in accord with the Helsinki Declaration of 1975. Approval for the study was granted by the NHS Research Ethics Committee London Camberwell St Giles. Consented participants were seen at the West London NHS Trust, where they underwent a range of clinical and cognitive assessments (Ritchie & Ritchie, 2012). The cohort comprised cognitively healthy volunteers aged 40-59 years. Here we examined baseline and follow-up data from the West London dataset. 210 individuals (62 male; 148 female) were tested at baseline, with 188 (89.5%) (55 male; 133 female) retained at two-years follow-up (Table 1). Mild cognitive impairment and dementia were ruled out based on detailed clinical assessment on each visit.

**Table 1.**
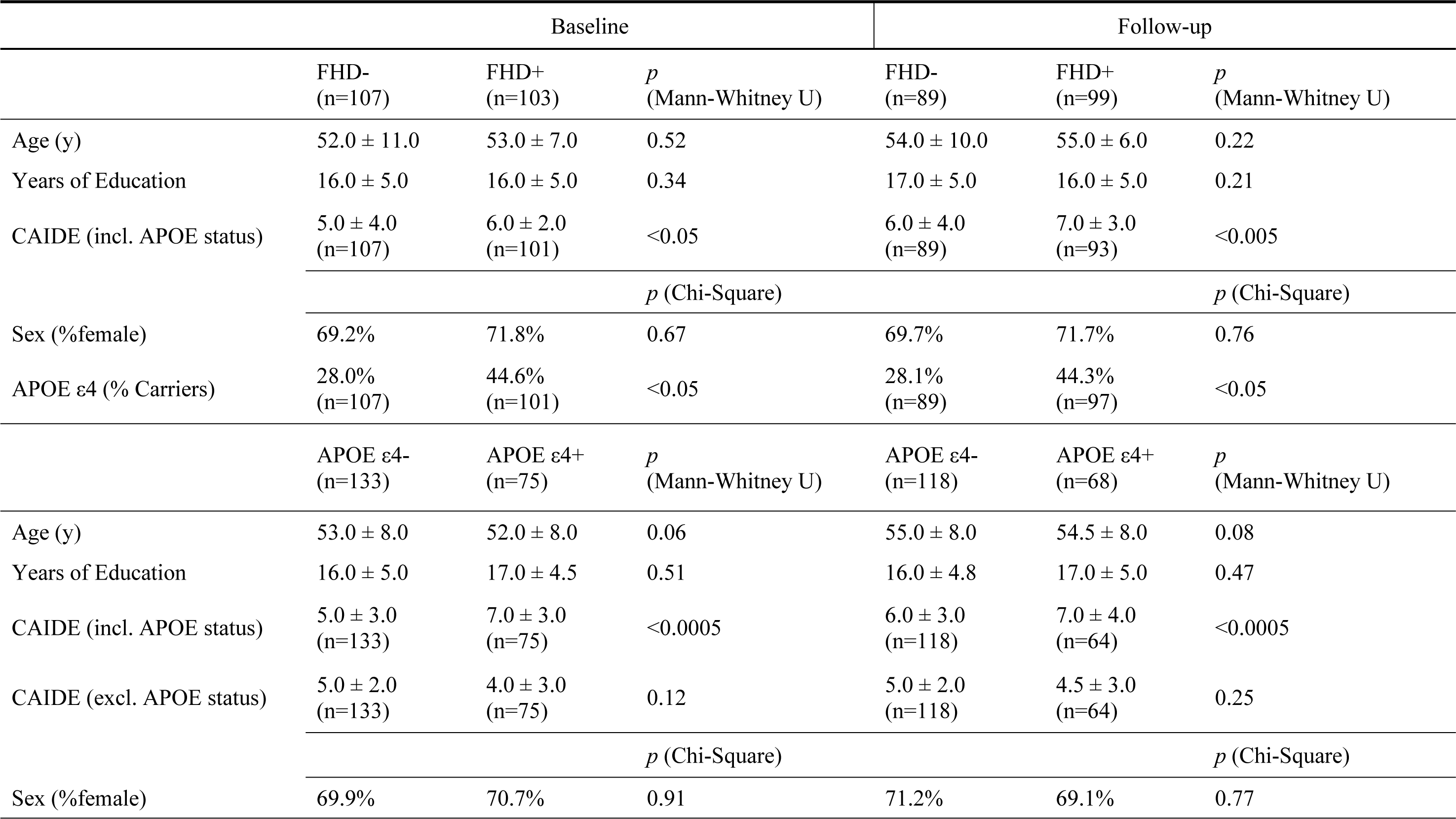
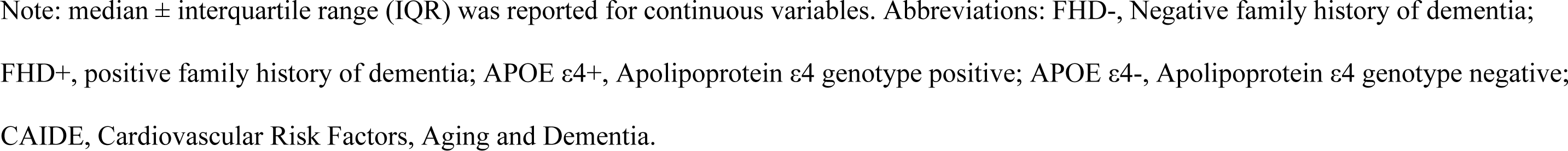
Demographic characteristic of the cohort at baseline and follow-up based on the dementia family history and on APOE genotype.

At baseline, 2 participants were missing APOE ε4 genotype information, and therefore the CAIDE scores. At follow-up, 6 participants were missing information for calculating their CAIDE scores, including the 2 missing APOE status. For cognitive tests, 2 participants were missing data at baseline, and 12 at follow-up. Therefore, the cognitive analyses cohort was N=208/N=176 at baseline/follow-up. 193 out of 210 participants finished MRI scanning at baseline, and 169 out of 188 at follow-up. Furthermore, at baseline, 6 were excluded due to incidental findings, and 58 due to not meeting the criterion of > 150 frames with minimal head motion (for details see the following section *2.4* *MRI data acquisition and processing*). At follow-up, 3 were excluded due to incidental findings and 73 due to < 150 frames with small head motion. The final fMRI analyses cohort was N = 128/N = 93 at baseline/follow-up. Please see the Supporting Information (SI) file and STable 1–2 and SFigure 1 for a description of any missing data.

**Figure 1.**
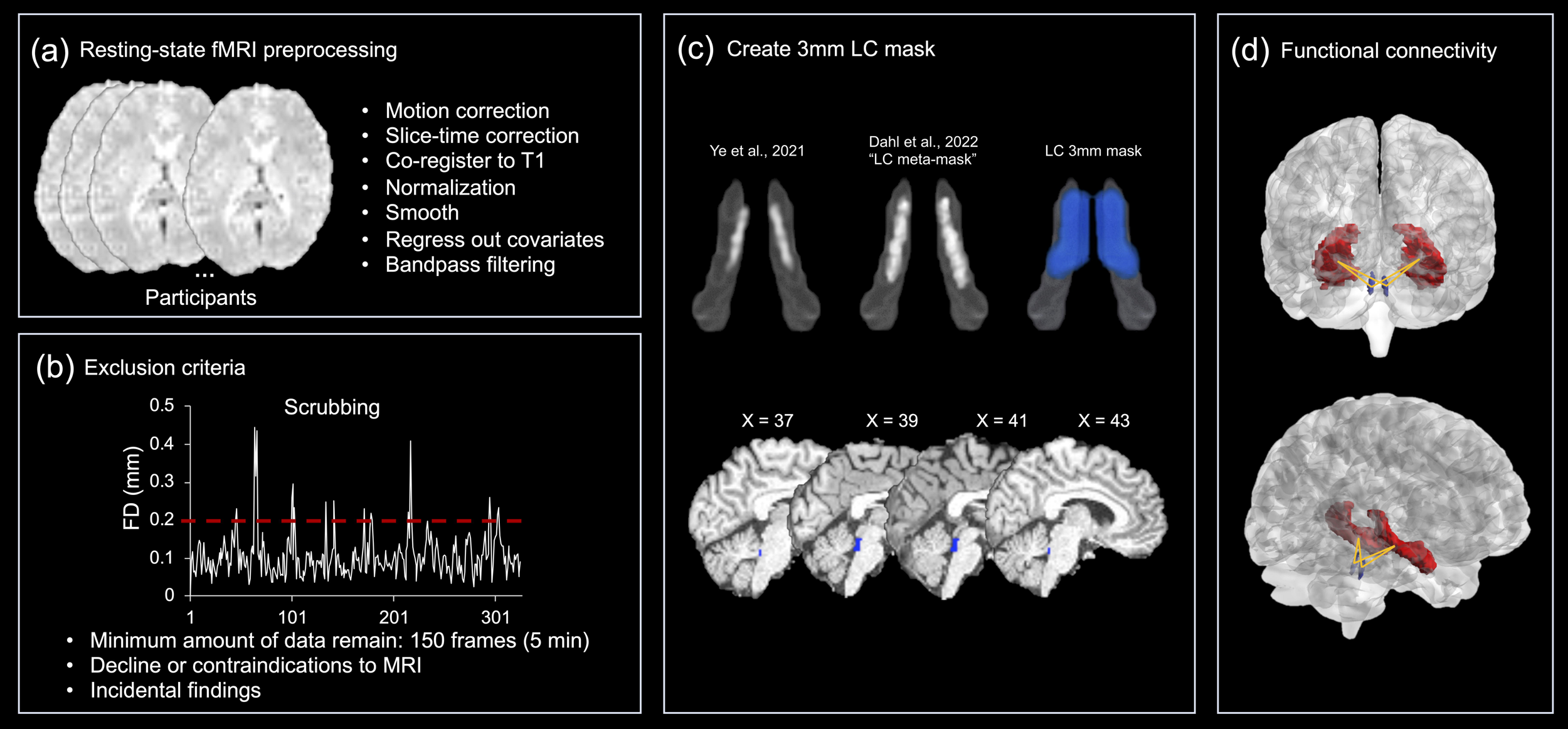
The flowchart of functional magnetic resonance imaging (fMRI) data processing. (a) The standard preprocessing pipeline for fMRI data. (b) Three exclusion criteria were used to exclude participants. A ‘scrubbing’ procedure was used to account for the effect of head movement on functional connectivity analyses (Power et al., 2012). (c) The top panels shows the study-specific 3mm locus coeruleus (LC) mask (in blue), which closely matched the 7T probabilistic atlas developed by Ye et al. (2021) and the LC meta-mask developed by Dahl et al. (2022). The bottom panel shows the study-specific mask superimposed on a participant’s T1w structural data in MNI space. (d) Functional connectivity was calculated between the bilateral LC (in blue) and the hippocampus (in red). The hippocampus was defined after *recon-all* pipeline using Freesurfer version 7.1.0.

### 2.2 Biological sex

While the dichotomies of sex and gender are no longer considered to be sharply discrete, in this study ‘sex’ was defined as an individual’s natal or biological sex and was self-reported in the Brain Injury Screening Questionnaire. The cohort is 70% female, reflecting higher female interest in the study, possibly due to the known female predisposition to developing Alzheimer’s disease.

### 2.3 Menopausal Status

Menopausal status was classified based on answers from a pregnancy and menstruation survey administered during the clinical assessments. Self-reported menopausal status was determined via answers to the question “Are you postmenopausal?”. Those who answered ‘Yes’ were categorized as postmenopausal, and those who answered ‘No’ were categorized as premenopausal. Of the 148/210 female participants, 55 (37.16%) were postmenopausal, 73 were premenopausal, and 20 who answered ‘don’t know’. Those who answered ‘Don’t Know’ (20/210) were excluded from further analyses.

### 2.4 Assessments

#### 2.4.1 Risk factors

##### APOE ε4 Genotyping

The process of APOE ε4 allele identification is outlined in detail in Ritchie et al. (2017). In brief, genomic DNA was isolated from blood samples and APOE genotyping was performed. All members of the research and clinical teams were blind to the result of APOE genotyping. In this study, APOE ε4 risk is determined by ≥1 APOE ε4 allele. 75/210 carried ≥1 APOE ε4 allele (See Table 1). The prevalence of APOE ε2ε4 in our cohort is 0.02% (see SFigure 2). Investigation of the effect of the APOE ε2ε4 allele can be found in the SI. The exclusion of APOE ε2ε4 carriers did not affect our main results and we therefore report results including APOE ε2ε4 carriers.

**Figure 2.**
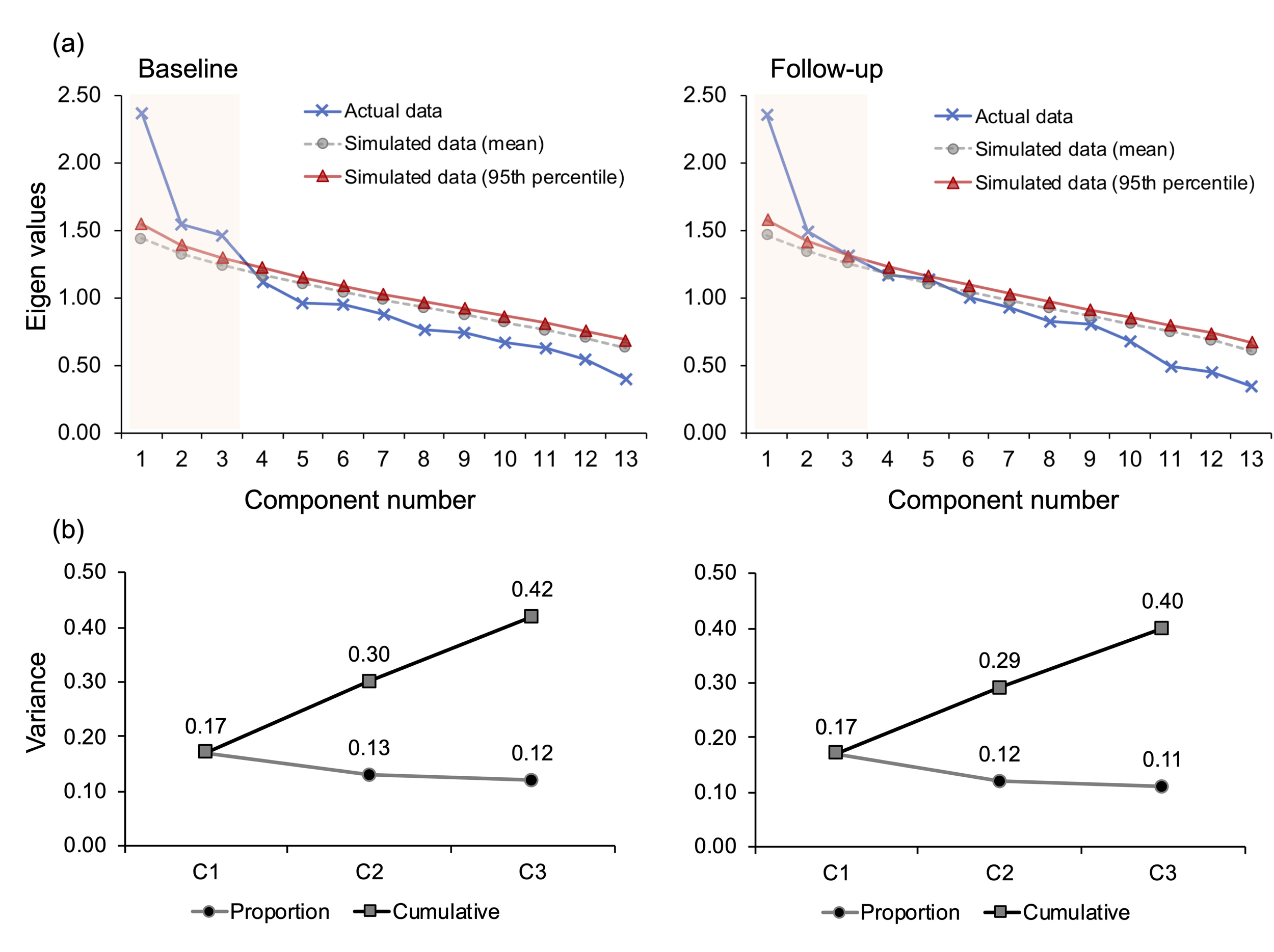
Data reduction of cognitive information. (a) Scree plots and parallel analysis for the baseline and follow-up datasets. Eigenvalues of the principal components obtained from the actual data (in blue line) were compared to those of random simulated data (mean values in grey line and 95th percentile in red line). Eigenvalues larger than 95th percentile of the randomly generated 500 eigenvalues (simulation) determined the number of components. (b) The three extracted components were rotated to be uncorrelated with each other. The proportion of variance explained by each component, and the cumulative variance explained by the three are presented for the baseline and follow-up datasets. Abbreviations: C1, component 1; C2, component 2; C3, component 3.

##### Family History of Dementia

FHD was determined by a ‘yes’/‘no’ question during clinical visits, where participants were asked whether a parent had a diagnosis of dementia. Participants were asked to include the dementia subtype if known, but answering ‘yes’ alone categorized a participant as FHD+. The answer ‘no’ likely captured both participants with no FHD, and participants for whom FHD was unknown. In summary, participants were defined as FHD+ if at least one parent was diagnosed with dementia. Cases where the FHD was unknown or partially known were not recorded outside of the binary yes/no scoring. 103/210 were FHD+.

##### Cardiovascular Risk Factors, Aging, and Incidence of Dementia (CAIDE) score

CAIDE is a composite scale of estimated future dementia risk based on mid-life cardiovascular measures (Fayosse et al., 2020; Sindi et al., 2015). It takes into consideration the individual’s age, sex, educational attainment, APOE ε4 genotype, activity level, BMI, cholesterol and systolic blood pressure (Kivipelto et al., 2006) and is scored on a range of 0 – 18. A higher score indicates greater risk. The CAIDE dementia risk score was calculated at baseline and follow-up.

#### 2.4.2 Cognitive testing

Cognitive function was assessed at baseline and follow up with the COGNITO neuropsychological battery (Ritchie et al., 2014), designed to examine information processing across a wide range of cognitive functions in adults of all ages and not restricted to those functions usually implicated in dementia detection in the older adults. Tests are administered using a tactile screen to capture information processing time as well as response accuracy and require about 40 minutes to complete. The tests, by order of presentation, are: reaction time; reading; comprehension of phonemes, phrases, and syntax; focused and divided attention in both visual and auditory modalities; visual working memory (visual tracking with auditory interference); the Stroop test; immediate, delayed, and recognition trials for verbal recall (name list); delayed recognition of spatial stimuli (faces); visuospatial associative learning; visuospatial span; form perception; denomination of common objects; spatial reasoning; copying of meaningful and meaningless figures; verbal fluency with se-mantic and phonetic prompts; immediate recall of a narrative; immediate recall of a description of the relative position of objects; vocabulary; implicit memory (recognition of new and previously learned material).

The COGNITO tests are designed to test several aspects of cognition, including attention (task: visual attention), memory (tasks: narrative recall, description recall, implicit memory, name-face association, working memory), language (tasks: phoneme comprehension, verbal fluency) and visuospatial abilities (task: geometric figure recognition) (Ritchie et al., 2014). Based on previous studies (Ritchie et al., 2021; Ritchie et al., 2018; Ritchie et al., 2017), 11 summary variables from the COGNITO battery capturing the above functions were used here (for a list see STable 2).

Additionally, we used the Visual Short-Term Memory Binding task (VSTMBT) (Parra et al., 2010), a computer-based task that assesses visual short-term memory binding of single features, e.g., complex shape or color combinations, or feature conjunctions, e.g., shape and color combinations. In the single feature condition, participants must identify whether the test stimuli (three random 6-sided polygons) are the “same” as or “different” to the studied stimuli in terms of shape (shape only) or color (color only). In the binding condition, participants are required to correctly identify if both the shape and color of the test stimuli match studied stimuli. The two summary variables from the VSTMBT were the percentage of correctly recognized items from the two conditions.

The Addenbrooke’s Cognitive Assessment (ACE) III (Hsieh et al., 2013) was recorded at the follow-up session, but not at the baseline. Analyses that focused on the subset of assessments performed at both testing sessions did not include the ACE III.

### 2.5 Behavioural data analyses Rotated principal component analysis

Rotated principal component analysis (rPCA), a dimensionality reduction technique, was adopted to cluster the above-mentioned 13 cognitive measures into related domains, reduce the number of multiple comparisons between the cognitive tests (Jolliffe & Cadima, 2016). This step maximized power to investigate the impact of risk factors on cognition and brain– behaviour relationships. rPCA was conducted by using the *psych* package (Version 2.0.12) in R software (https://www.r-project.org/), following steps as: (1) *Preprocessing*. Listwise deletion procedure was used to deal with the missing data. Only participants who had no missing observations were kept (N = 208 at baseline; N = 176 at follow-up). (2) *Component extraction and estimation*. Principal component analysis was used to extract a smaller number of components that capture the most variance of the initial variables, representing the common cognitive processes underlying the 13 original cognitive measures. Scree plots and parallel analysis were used to determine the number of components (Horn, 1965). Parallel analysis generated a set of random correlation matrices (n = 500) by using the same number of variables (n = 13) and participants (N = 208 at baseline; N = 176 at follow-up). Then, we compared the solution to the random simulated normal data. The eigenvalues obtained from the actual matrix were compared with those obtained from the randomly generated matrices, and the estimated number of components was determined by selecting components with eigenvalues larger than 95th percentile of the randomly generated components’ eigenvalues (See also SI). (3) *Rotation*. To examine the component structure among the initial variables, an orthogonal rotation method Varimax that constrains the components to be uncorrelated, was applied to the coefficient matrix (Kaiser, 1958). (4) *Component coefficients*. Component coefficients (*r*), reflecting the correlation between each of the components and each of the original variables, were thus obtained. Given the sample size in this study (N > 200), component coefficients that were > 0.4 were deemed practically significant (Hair Jr. et al., 1998; Stevens, 1992). (5) *Component scores*. Component scores were calculated by a regression method where the regression weights were found from the inverse of the correlation matrix times the coefficient of each variable on the corresponding components. The coefficients were used to interpret the components, and component scores were used for the following analyses. (6) *Component similarities*. Components were extracted at the baseline and follow-up respectively. To test the similarity of components extracted at each study session, to be capturing similar cognitive functions, Tucker’s congruence coefficient was calculated (Lorenzo-Seva & ten Berge, 2006). In subsequent analyses, we used these cognitive components, rather than the individual tests to measure the impact of risk factors on cognition and brain–behaviour relationships.

### 2.6 MRI data acquisition and processing

Imaging data were obtained as part of a multimodal examinations in a 3T Siemens Verio MRI scanner and with 32-channel head coil (https://preventdementia.co.uk/for-researchers/). Resting-state fMRI data were acquired with T2*-weighted echo-planar imaging (EPI) sequence. Total acquisition time for the resting state scan was 11 minutes 06 sec. The eyes were closed during rest. 330 volumes were acquired, and each volume contained 35 slices (interleaved acquisition), with slice thickness of 3 mm (repetition time (TR) = 2000ms, echo time (TE) = 30ms, flip angle (FA) = 80o, Field of View (FOV) = 192 × 192mm2, voxel size = 3 mm3 isotropic). A 3D T1-weighted magnetization prepared rapid gradient-echo image (MPRAGE, 160 slices, voxel size = 1 mm3 isotropic, TR = 2300ms, TE = 2.98ms, FOV = 240 × 256mm2, FA = 9o) was also acquired. All scans were repeated after approximately two years on the same scanner using the same protocol.

Standard preprocessing procedures for resting-state fMRI data were performed with SPM12 (Statistical Parametric Mapping, https://www.fil.ion.ucl.ac.uk/spm/software/spm12/) and the AA pipeline software (Cusack et al., 2015) implemented in MATLAB R2019a (The MathWorks, United States). We discarded the first 5 volumes to allow for adaptation to the environment and equilibrium of the MR signal. In this pipeline (Figure 1a), we performed slice timing correction, motion correction, co-registration of functional and structural images, normalization into standard MNI space, and spatial smoothing. Spatial normalization was performed using SPM12’s segment-and-normalize procedure, whereby the T1 structural was segmented into grey matter (GM), white matter (WM) and cerebrospinal fluid (CSF) and normalized to a segmented MNI-152 template. These normalization parameters were then applied to all EPIs. The quality of spatial normalization was visually inspected for each participant and no participants showed a normalization failure. The data were then smoothed with a Gaussian kernel of 6mm full width at half maximum. We also applied a general linear model (GLM) that includes 24 head motion parameters – 6 rigid-body motion parameters, the first-order temporal derivative of them and quadratic terms of the original parameters and their derivatives (Satterthwaite et al., 2013) – and averaged signals from WM and CSF as covariates to reduce any residual effects of head motion and physiological noise. Lastly, all preprocessed data were temporally band-pass filtered (0.01-0.08 Hz) to remove low-frequency drift and high-frequency physiological noise (Salvador et al., 2008; Zuo et al., 2010).

Because brain signal from the brainstem is susceptible to head movements, we performed careful measurement and control for head motion, by using a “scrubbing” procedure (Power et al., 2012). We first calculated an index of data quality called framewise displacement (FD) (for details please also see SI), which measures how much the head has changed position from one frame to the next, and can therefore be used to flag frames of suspect quality based on the threshold of FD > 0.2 mm, in order to remove motion-contaminated data before calculating functional connectivity (Power et al., 2012). A participant inclusion criterion was then set to ensure a minimum amount of data remained after scrubbing, i.e., at least 150 frames (∼5 min) of data (Figure 1b), based on wide agreement that ∼5 min of fMRI data is an adequate for resting state functional connectivity analyses (Van Dijk et al., 2010). After completing the scrubbing procedure, the included participants had 74.0% ± 14.8% frames at baseline and 73.5% ± 15.0% at follow-up. Finally, we calculated the mean FD across the remaining good frames, which was further regressed in the group-level analyses (Power et al., 2014). The mean FD was 0.12 ± 0.01 at baseline and 0.12 ± 0.01 at follow-up. There were no significant associations of mean FD with any of the risk factors or the LC–Hippocampus functional connectivity (See SI). The fMRI-based exclusion criteria are also illustrated in Figure 1b.

To ensure high accuracy in the localization of the LC, we created a binary LC mask of 3 mm3 isotropic voxel size that closely matched with two previously published LC maps (Dahl et al., 2022; Ye et al., 2021), which were developed in the standard MNI space with 0.5 mm3 isotropic voxel size. In order to achieve the highest precision, instead of resampling the LC maps to match the fMRI data’s voxel size (3 mm3) using automatic scripts, we manually drew a 3 mm3 binary LC mask in MNI space with reference to the two LC maps. This is because the automatic scripts did not perform well when resampling the two referenced LC maps from high to low resolution due to the small size of these two LC regions of interest. The FSL software (https://fsl.fmrib.ox.ac.uk/fsl/fslwiki/) was used for the drawing process, by following the same procedure as in Plini et al. (2021). The two 0.5 mm3 LC maps were superimposed on a blank 3 mm3 template in standard MNI space. The drawing process was then performed on this blank template by adding or removing voxels to include different anatomical locations of the two referenced maps without crossing the dorsal and medial raphe nuclei, as defined by Beliveau et al. (2015), and the cerebellar vermis white matter. At the end of this procedure, the new bilateral LC mask was symmetrical and consisted of 16 3 mm3 voxels, was anatomically inclusive of the 7T LC map (Ye et al., 2021) and the LC meta-map (Dahl et al., 2022), and did not encroach on other pontine nuclei (Figure 1c).

In an independent study (Dounavi et al., 2022), T1-weighted MPRAGE of this cohort were corrected for field inhomogeneities using the Advanced Normalisation Toolbox (ANTs) N4 algorithm (Tustison et al., 2010) and processed using Freesurfer version 7.1.0 (Desikan et al., 2006). The *recon-all* pipeline was run for every participant with standard settings. The brain masks and surfaces were inspected following recon-all and manual corrections were applied: (a) in the form of erosion of non-brain voxels from the brain mask or non-WM voxels from the WM mask, (b) in the form of filling of areas where the brain was not correctly identified, or (c) with the addition of control points in cases where white matter was not successfully identified. Following these steps, we generated individual masks of the hippocampus, which were then applied to the fMRI data to extract the time course of the region (Figure 1d). The functional connectivity (FC) between the LC and the hippocampus was derived using Pearson’s correlation coefficient (*r*) on the denoised time courses (Figure 1d). The *r* value was then transformed to z value using Fisher’s *r*-to-*z* transformation to improve normality for further statistical analysis. To avoid the formation of artificial anti-correlations (Anderson et al., 2011; Murphy et al., 2009), we did not perform global signal regression. In addition, volume of the hippocampus (HCV) was derived for each participant at baseline and follow-up as a covariate for functional connectivity analyses.

### 2.7 Statistical approach

All statistical analyses were performed in R software. The normality of the data was assessed by combining the visualization of a quantile-quantile plot and the Shapiro–Wilk test. Demographic and clinical information of the study cohort was analyzed across risk groups using chi-square (*χ2* tests) for categorical variables and Mann Whitney U tests for continuous (discrete) variables, given that they were not normally distributed in this cohort.

To investigate the cross-sectional effects of risk factors (APOE ε4 genotype and FHD) on cognition and brain function we used multiple linear regression models. Age, years of education were included as covariates for cognition models, and mean FD values and HCV were additionally included as a covariate for models of functional connectivity. The effects of APOE ε4 and CAIDE risk factors were modelled independently to avoid modelling the variance associated with APOE genotype in the same model twice. Spearman correlation analyses were used to assess the associations between cognition and CAIDE score given the non-normal distribution of CAIDE in our cohort. For the CAIDE model, we did not control for the age, years of education, as they are considered in the CAIDE scoring.

For any observed risk effect on cross-sectional changes of cognition or functional connectivity, interactions between risk factors and the LC–Hippocampus functional connectivity on cognition were assessed through multiple regression models with the cognition as the dependent variable, the LC–Hippocampus FC, risk factor and the risk × FC interaction as independent variables, and age, years of education, and HCV as covariates. Age, sex, and years of education were not included as covariates when CAIDE was included in the models, because CAIDE includes these variables in its scoring. Biological sex was investigated as variables of interest in these analyses especially given the higher proportion of females in the sample. If a sex effect was found, any interactions with risk factors, and the effect of menopausal status was also considered because the cohort age range [52+/- 11 years] cover menopause transition in females, which is known to affect cardiovascular health and cognition.

For any observed interactions, we plotted the regression of LC–Hippocampus FC on cognitive performance for each level/value of the risk factor, to interpret the effect (Aiken & West, 1991). For the continuous variable CAIDE, we applied the Johnson-Neyman technique (Bauer & Curran, 2005; Johnson & Fay, 1950), to test the significance of the slopes of the simple regression lines on each value of the CAIDE scores (ranged from 0-18). This examined the CAIDE interval at which the brain-cognition coupling was significant. All continuous variables in the interaction terms, i.e., CAIDE and FC, in the moderation models were mean-centered (subtracting each the mean from each value) to avoid multicollinearity.

Longitudinal changes were first assessed using random intercept linear mixed effect models (Cnaan et al., 1997) for the entire cohort. Specifically, we evaluated longitudinal changes of cognition (Eq. 1) and functional connectivity (Eq. 2) over two years.

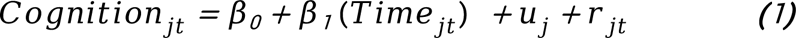

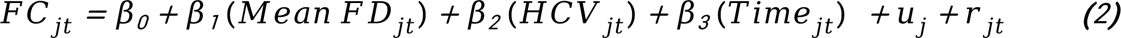

For each participant *j*, measures of cognition/functional connectivity at each time point *t* served as dependent variables, and the timepoint (a categorical variable with two levels: baseline and follow-up) served as the independent variable. Mean FD and HCV at each time point *t* were included as covariates for the functional connectivity model only (Eq. 2). *u_j_* represented individual variability in the intercepts, i.e., a random effect.

The presence of a longitudinal change for the entire cohort was then further assessed to determine whether it varied in relation to the risk factors, as assessed by the interaction term (*Risk_j_* × *Time_jt_*) Age at baseline, sex, and years of education, were included as covariates for the cognition models that did not include CAIDE (Eq. 3). Mean FD and HCV were additionally included as covariates for the functional connectivity models (Eq. 4).

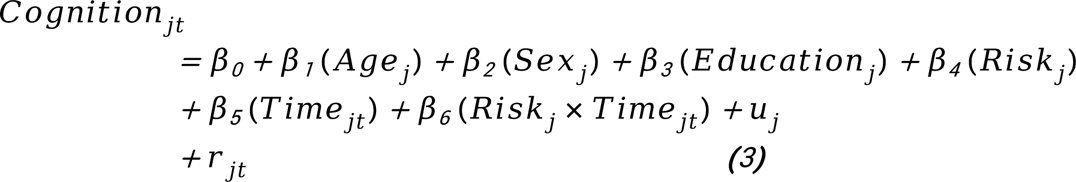

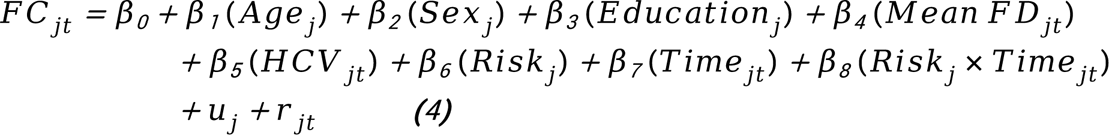

Lastly, the presence of risk related longitudinal change in cognition or FC was further investigated to determine whether risk factors influence brain-cognition association longitudinally. Multiple linear regression modelling was used with a percentage change score [i.e. (follow-up - baseline)/baseline] in cognition as the dependent variable, and a risk factor, a percentage change in FC, and their interaction as independent variables. Baseline age, sex, years of education and changes in HCV were included as covariates. Only changes in HCV were controlled for when assessing CAIDE in the models.

Although the baseline and follow-up datasets were not independent, we considered each in its own right, in addition to testing for longitudinal changes in cognition and functional connectivity. During the two years, a proportion of our participants may have sustained brain health that are yet subthreshold to clinical manifestations, or that may not yet manifest as longitudinal change. Statistical results were corrected for multiple comparisons.

## 3. Results

### 3.1 Demographic characteristics

Demographic characteristics of the cohort at baseline and follow-up, stratified by APOE ε4 genotype and FHD, are shown in Table 1. There were no significant differences in age, sex or years of education between the groups. APOE ε4 allele genotype was more frequently found in the FHD+ than FHD-group at baseline (*p* = 0.01) and follow up (*p* = 0.02). CAIDE scores (including APOE status) were significantly higher for the FHD+ than FHD-group at baseline (*p* = 0.03) and follow up (*p* = 0.003). Naturally, CAIDE scores including APOE status were significantly higher for the APOE ε4+ than APOE ε4-group at baseline (*p* = 0.0003) and follow up (*p* = 0.0002), but when APOE status was excluded the CAIDE scores did not differ between the two groups (Table 1).

### 3.2 Cognitive domains

We first performed parallel analyses and plotted scree plots to determine the number of components that best represented the original 13 cognitive measures at baseline and follow-up, separately. Results showed that eigenvalues of the first three components were larger than 95th percentile of the randomly generated eigenvalues at both study timepoints (Figure 2a), which indicated that the three component (C) solution best represented the data. The three components were then rotated to be uncorrelated with each other, and could cumulatively explain a total of 41% percentage of the variance, at baseline (C1 = 17%; C2 = 13%; C3 = 12%) and 40% at follow-up (C1 = 17%; C2 = 12%; C3 = 11%) (Figure 2b).

The loading values (weights) of the cognitive measures (Figure 3a) reflect the relationships between the original measures and the corresponding components. Measures with higher coefficients were more closely related to the components. Based on the cognitive functions that the highest loading measures tapped into (for details see STable 2), in subsequent analyses we refer to: C1 as ‘episodic and relational memory’; C2 as ‘working and short-term (single-feature) memory’; C3 as ‘verbal, visuospatial functions, and short-term (conjunctive) memory.’

**Figure 3.**
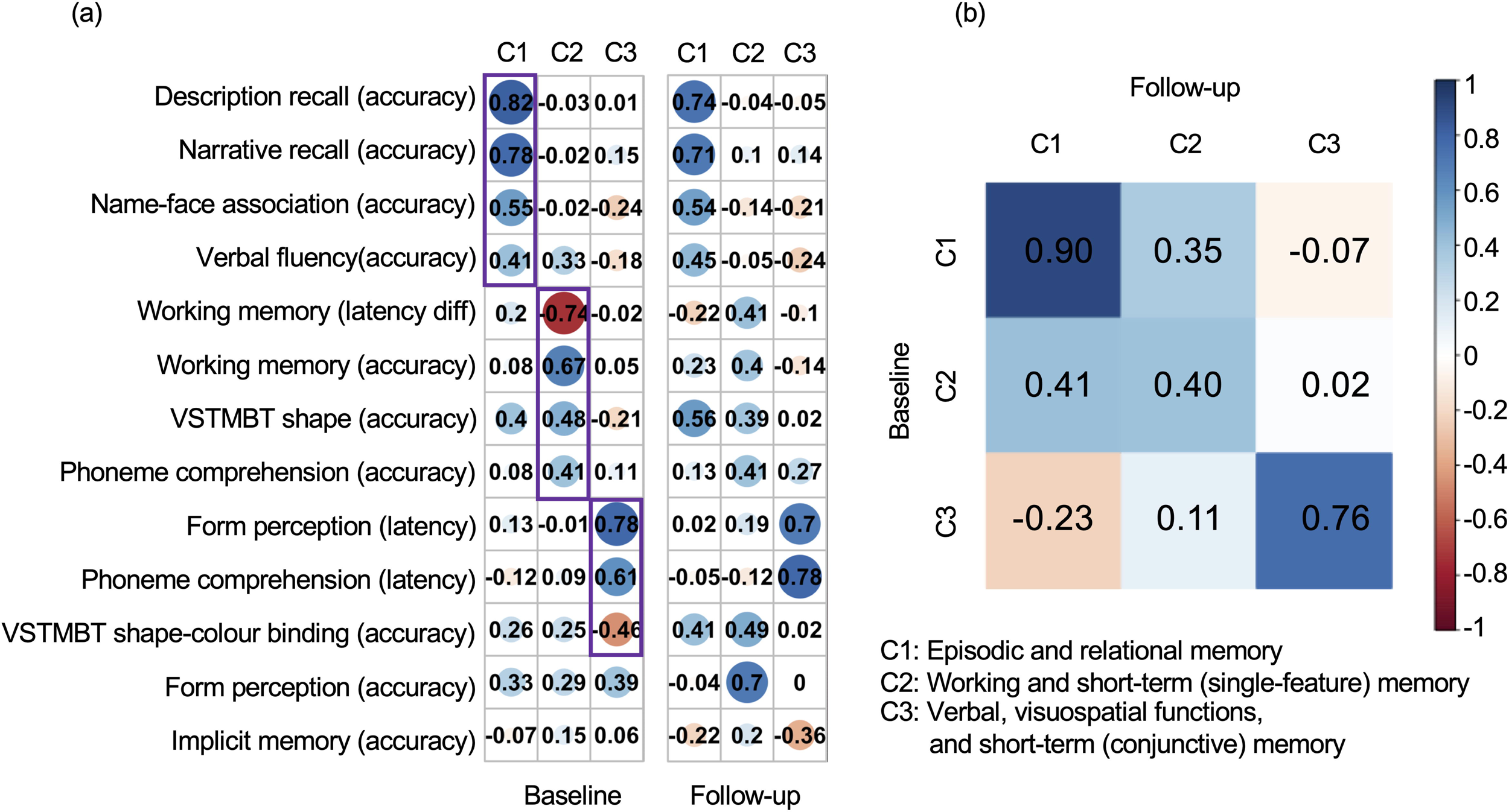
Interpretation of data-derived cognitive domains. Component (C) coefficients and Tucker’s congruence coefficients between (a) baseline and (b) follow-up, are shown. (a) The coefficients for 13 original cognitive measures (in rows) on the three components (in columns). A larger absolute coefficient (darker colors and larger solid circles) represents a closer relationship between the cognitive measure and corresponding component. Cool/warm colors represent the positive/negative relationships between cognitive measures and components. Only cognitive measures with a coefficient > 0.4 on each component, highlighted with purple rectangles, were used to interpret the cognitive functions driving each component. (b) Similarities among the components across baseline (in rows) and follow-up (in columns) were measured by Tucker’s congruence coefficients. The diagonal values represented the similarities for each of the components with itself across time, and off diagonal values indicated the similarities for each of the components with the other two components across time. Cool/warm colors represent the positive/negative relationships with darker colors representing correlation magnitude, as shown in the color-bar scale. Abbreviations: VSTMBT, visual short-term memory binding test; diff, difference.

The highest loading measures and their coefficients for each component differed for the cross-sectional and longitudinal data. For example, verbal functions, visuospatial functions, and short-term (conjunctive) memory loaded most strongly on the third component at baseline. At follow-up, implicit memory rather than short-term memory loaded on the third component (Figure 3a). We found high similarity between baseline and follow-up for C1 (ϕ = 0.90), and C3 (ϕ = 0.76), with relatively low similarly for C2 (ϕ = 0.40) (Figure 3b).

### 3.3 Impact of risk factors on cognition

#### Episodic and relational memory

At baseline, the multiple linear regression model showed a significant positive association between episodic and relational memory with APOE ε4 allele [*β* (*SE*) = 0.28 (0.13), *p* < 0.05], independently of sex, age and years of education (Table 2). At follow-up, there was a trend effect of APOE ε4 allele [*β* (*SE*) = 0.27 (0.15), *p* = 0.07]. Education was significantly positively associated with this domain at baseline [*β* (*SE*) = 0.09 (0.02), *p* < 0.0001] and follow-up [*β* (*SE*) = 0.10 (0.02), *p* < 0.0001]. Sex did not interact with the APOE ε4 allele in its effect on episodic and relational memory. The baseline effect of APEO ε4 on episodic and relational memory became a trend effect (SFigure 6) when corrected for multiple comparisons and needs to be interpreted with caution.

**Table 2.**
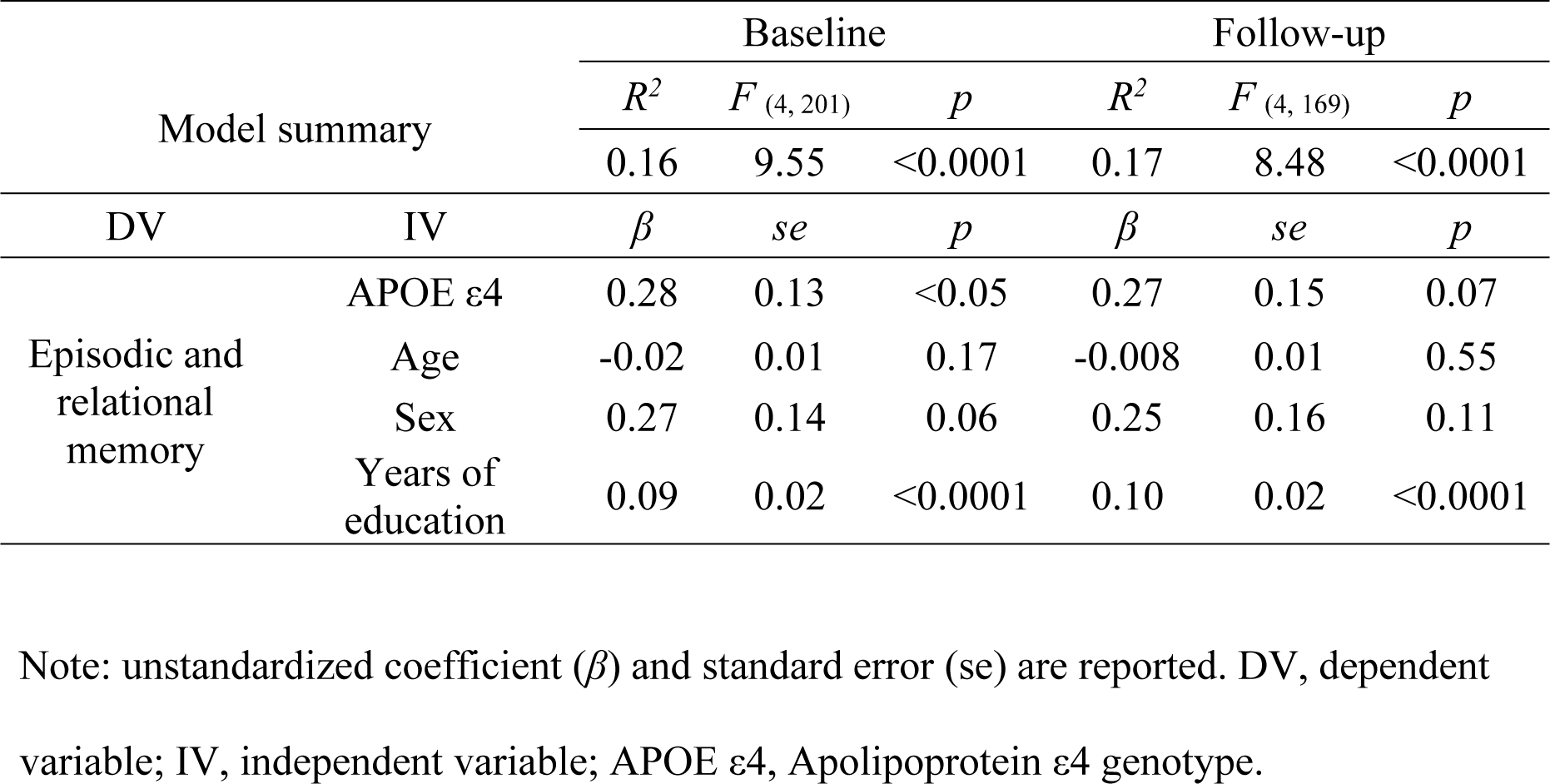
Statistical summary of the multiple regression model testing the relationship between APOE and episodic and relational memory.

The multiple linear regression models showed no associations between FHD and episodic and relational memory, at either timepoint. Spearman correlation analyses showed no significant associations between the CAIDE score and episodic and relational memory, at either timepoints.

#### Working and short-term (single-feature) memory

We found no associations between APOE ε4, FHD, or CAIDE score with working and short-term (single-feature) memory. Education was significantly positively associated with performance in this domain, independently of the other factors, at baseline [*β* (*SE*) = 0.04 (0.02), *p* < 0.05] only.

#### Verbal, visuospatial functions, and short-term (conjunctive) memory

We found no associations between APOE ε4 or FHD with verbal, visuospatial functions, and short-term (conjunctive) memory. Age was significantly negatively associated with performance in this domain, independently of the other factors, at baseline [*β* (*SE*) = -0.04 (0.01), *p* < 0.005]. The CAIDE score was significantly associated with verbal, visuospatial functions, and short-term (conjunctive) memory at baseline (*rho* = -0.17, *p* < 0.05, Figure 4a), and at follow-up (*rho* = -0.20, *p* < 0.01, Figure 4b). Higher CAIDE scores were significantly associated with worse performance.

**Figure 4.**
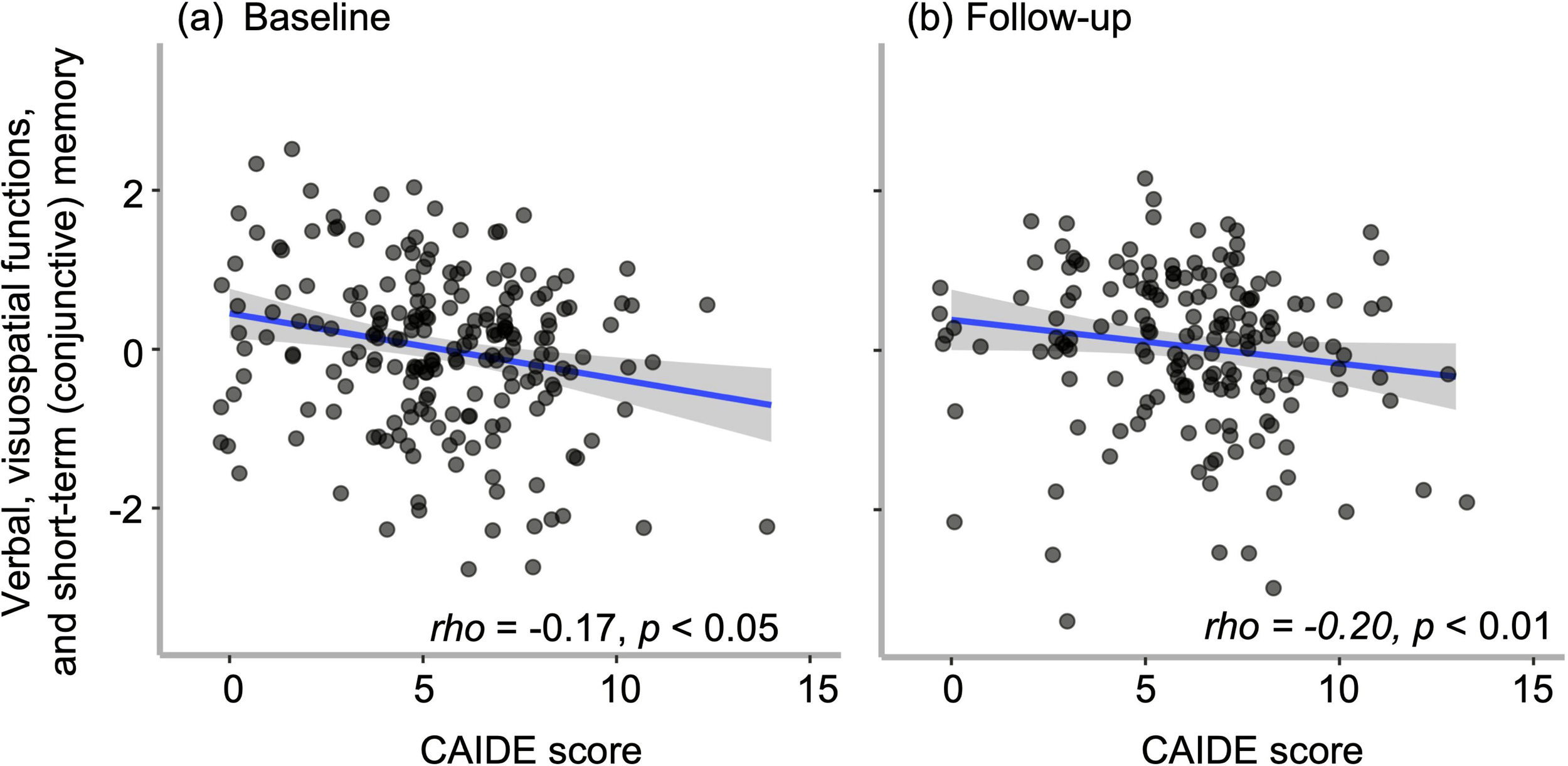
The associations between the CAIDE dementia risk score and verbal, visuospatial functions, and short-term (conjunctive) memory at (a) baseline and (b) follow-up. Abbreviation: CAIDE, Cardiovascular risk factor, Aging, and Incidence of Dementia.

As the cohort was 70% female, and the age range (40-60 years) covers the menopausal transition, which is known to affect cardiovascular health and cognition, the effect menopausal status on the relationship between CAIDE score and cognition was investigated. The CAIDE score includes sex in its calculation, with ‘male’ scored higher. Therefore, the effect of sex on relationship between the CAIDE risk and cognition not investigated. However, there was a significant interaction between CAIDE and menopause on verbal, visuospatial functions and short-term (conjunctive) memory at baseline [*β* (*SE*) = -0.20 (0.09), *p* < 0.05] and at follow-up [*β* (*SE*) = -0.29 (0.13), *p* < 0.05] (Table 3). Increasing CAIDE was associated with worse cognitive performance in post-menopausal females, but not in pre-menopausal counterparts. This effect was controlled for age effects, as post-menopausal women were overall older than pre-menopausal counterparts (post-: median ± IQR: 49.0 ± 8.0 vs pre-: 56.0 ± 4.0).

**Table 3.**
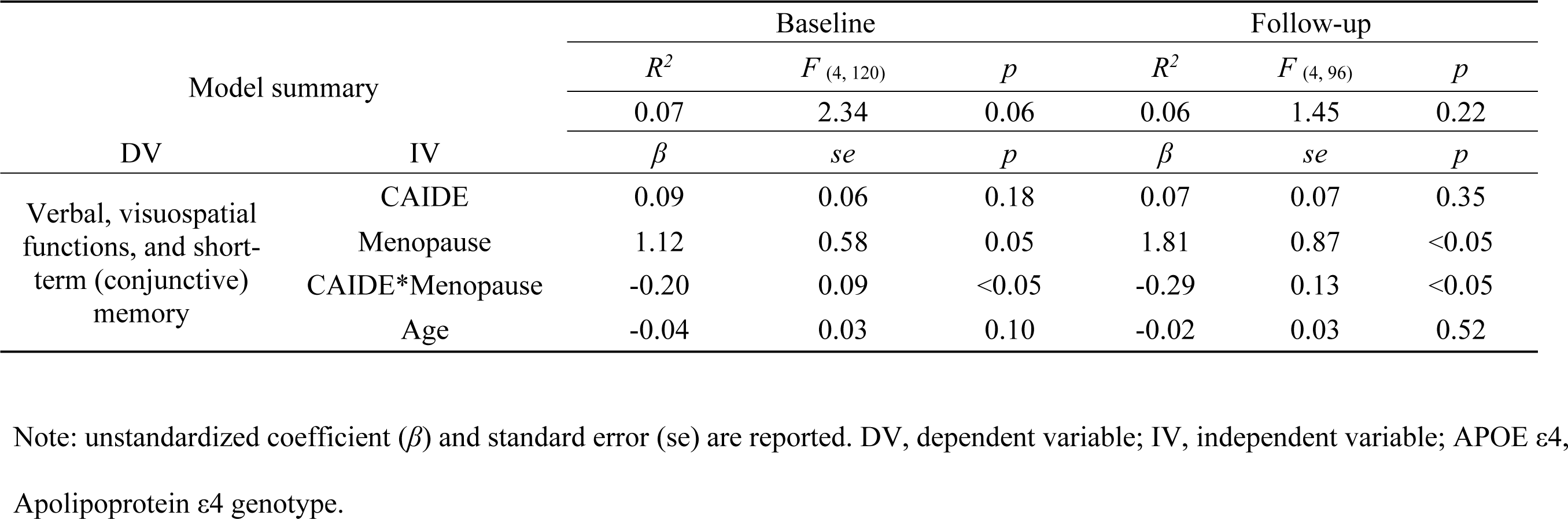
Statistical summary of the multiple regression model testing the interaction between CAIDE and menopausal status.

There was no significant longitudinal change of cognition in any of the three cognitive domains over two years.

### 3.4 Longitudinal change of LC–Hippocampus functional connectivity

We found a significant negative association between time and the LC–Hippocampus functional connectivity [*β* (*SE*) = -0.06 (0.03), *p* < 0.05], after controlling for mean FD and HCV (Table 4), indicating reduced connectivity at two-years follow-up relative to baseline across the whole cohort (Figure 5). Sex did not interact with time in affecting the LC– Hippocampus functional connectivity.

**Table 4.**
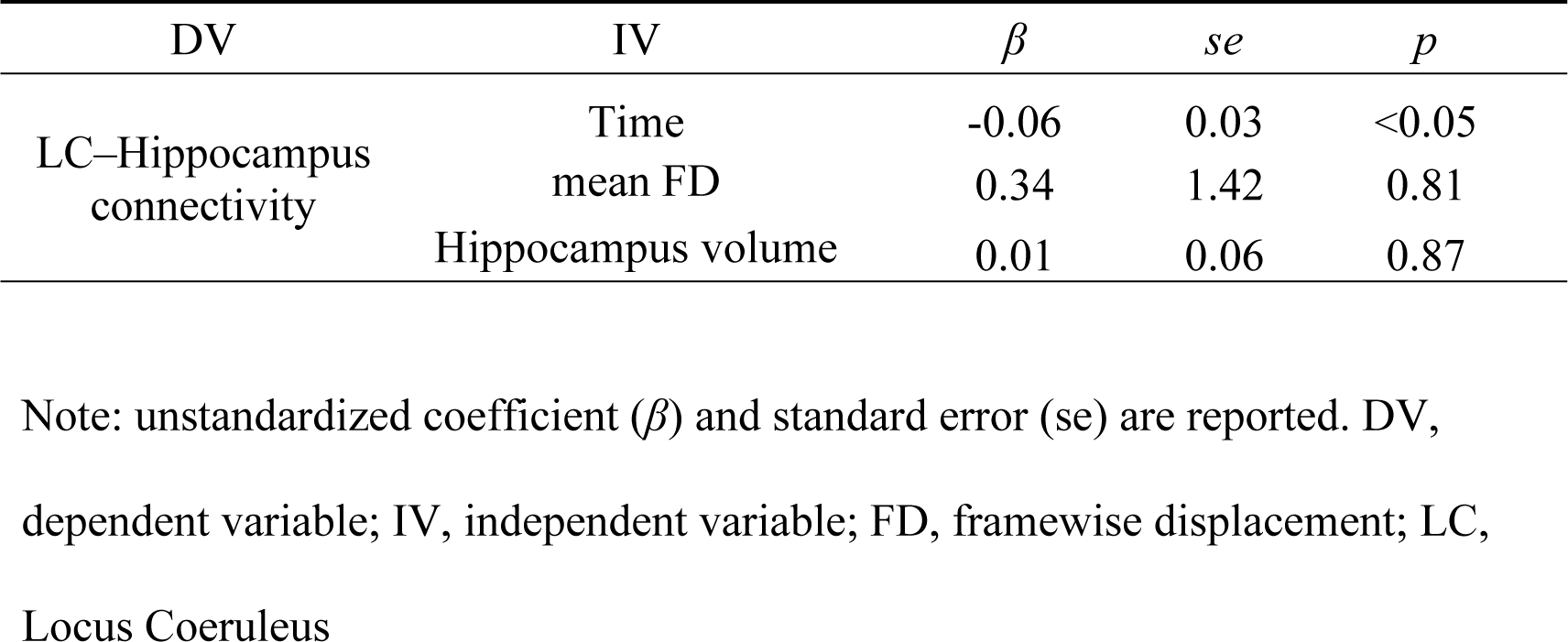
Statistical summary of longitudinal linear mixed effect model testing for longitudinal change in the LC–Hippocampus functional connectivity.

**Figure 5.**
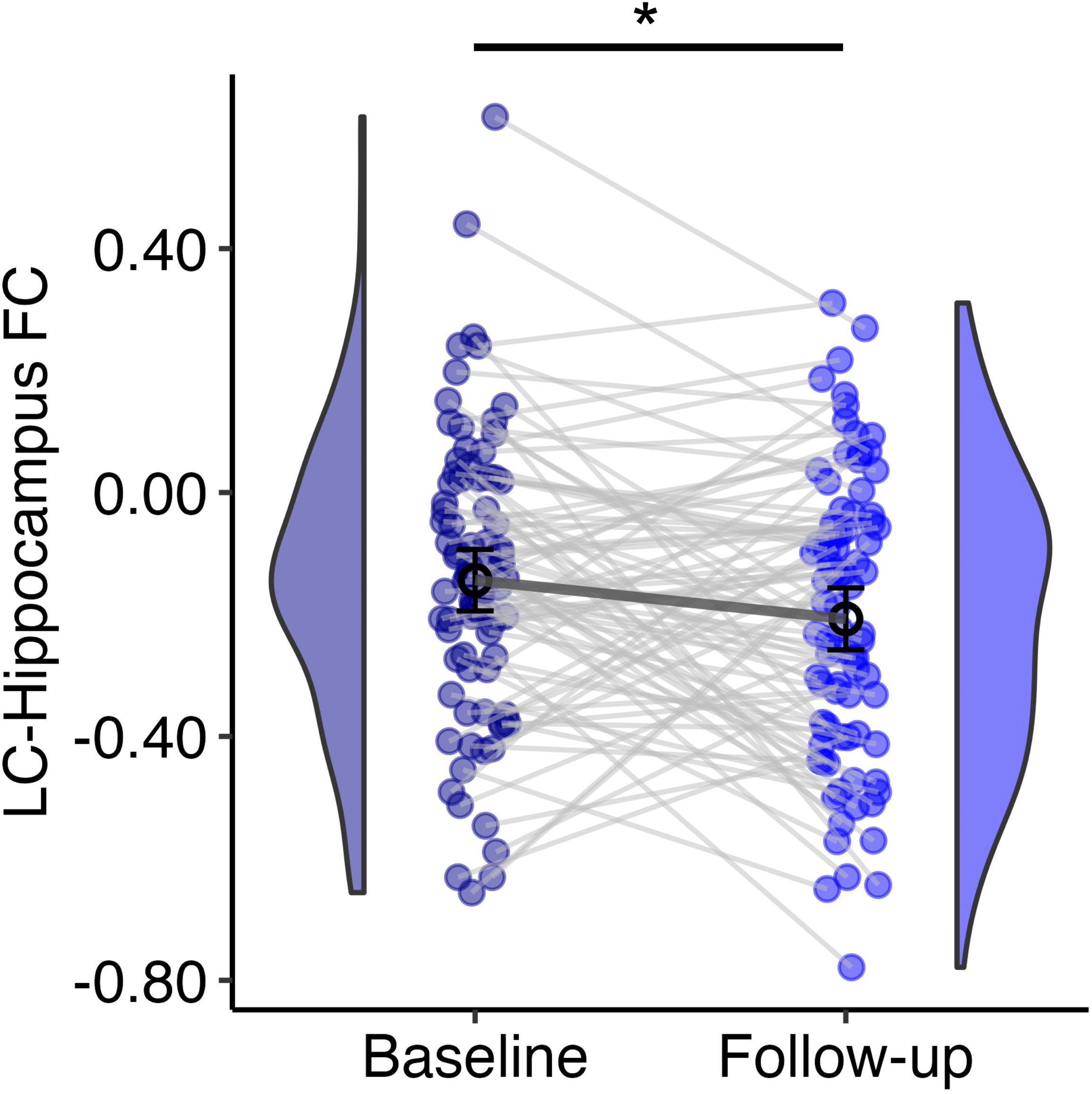
Longitudinal decline of the LC–Hippocampus functional connectivity over 2 years in cognitively healthy middle-aged adults. * = *p*<0.05. Abbreviations: LC, Locus Coeruleus.

### 3.5 Impact of risk factors on the LC–Hippocampus functional connectivity and brain– behavior relationships

There were no significant associations of *APOE* ε4, FHD, or CAIDE scores with the LC– Hippocampus functional connectivity, cross-sectionally, or longitudinally. At baseline, none of the risk factors moderated the relationship between functional connectivity and cognition. At follow-up, the multiple linear regression model with CAIDE, LC–Hippocampus FC, and CAIDE × LC–Hippocampus FC interaction term as independent variables, and verbal, visuospatial functions, and short-term (conjunctive) memory as the dependent variable showed a significant negative association between the interaction term (CAIDE × LC–Hippocampus FC) and verbal, visuospatial functions, and short-term (conjunctive) memory [*β* (*SE*) = -0.54 (0.22), *p* < 0.05] (Table 5). In individuals with low/high CAIDE scores, higher functional connectivity was associated with better/worse cognition (Figure 6). Menopausal status did not have an effect on this relationship.

**Table 5.**
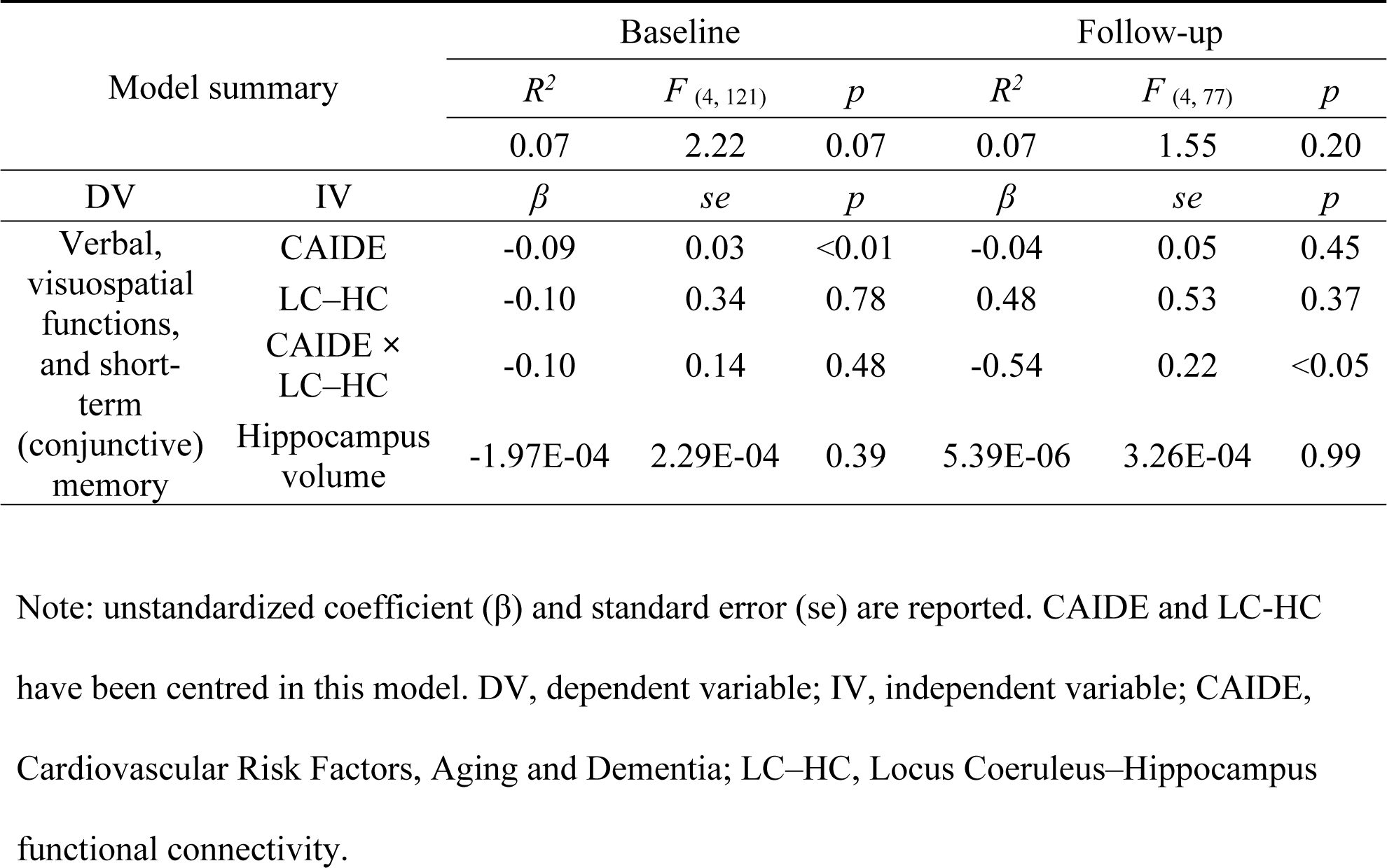
Regression coefficients of the model testing the relationship between CAIDE, LC– Hippocampus and verbal, visuospatial functions, and short-term (conjunctive) memory.

**Figure 6.**
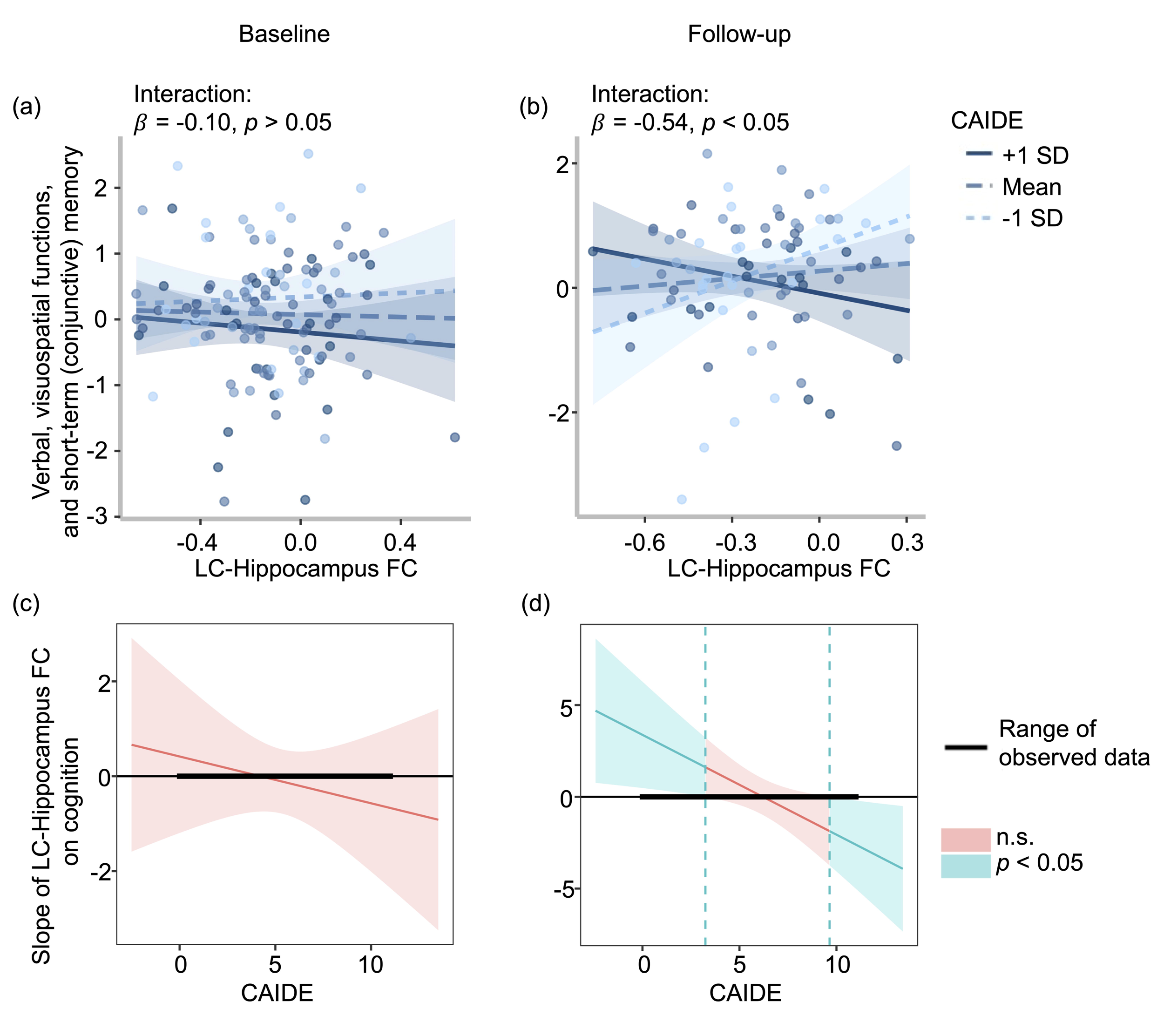
The CAIDE score significantly moderated the brain–behaviour relationship. The relationship was plotted at the fixed values of CAIDE (mean and ± 1 SD) to show the effect of CAIDE at (a) baseline and (b) follow-up. The LC–Hippocampus functional connectivity and the verbal, visuospatial functions, and short-term (conjunctive) memory were positively associated for individuals with low CAIDE score, and negatively associated for those with high scores, at (b) follow-up, but not baseline (a). Johnson-Neyman plots show the specific range of CAIDE scores where FC was significantly associated with cognition at (c) baseline and (d) follow-up. The bolded horizontal line shows the range of CAIDE scores of our cohort. Two blue dashed vertical lines indicate the interval of significant (blue area) and non-significant (pink area) brain-behaviour relationships. At follow-up, there was a significant positive relationship between FC and cognition for individuals with a CAIDE score < 3.25 and a significant negative relationship for individuals with a CAIDE score > 9.65. Abbreviations: LC, locus coeruleus; CAIDE, Cardiovascular risk factor, Aging, and Incidence of Dementia; FC, functional connectivity; SD, standard deviation; n.s., not significant.

## 4. Discussion

It is well acknowledged that AD neuropathology starts decades before clinical manifestations, but the brain mechanism of sporadic AD and its interactions with risk factors in midlife remain unclear. We examined whether risk factors for late-life AD were associated with cognition, functional connectivity between two of the key structures in AD neurogenesis – the LC and the hippocampus – and the brain-behaviour relationship between them, cross-sectionally and longitudinally in a cohort of middle-aged, cognitively healthy individuals. We found that the CAIDE scores were significantly negatively associated with verbal, visuospatial functions and short-term (conjunctive) memory functions in this cohort. Menopausal status significantly interacted with CAIDE on cognition, independently of age. Post-menopausal women showed worse cognition with increasing CAIDE scores. Furthermore, the CAIDE score significantly moderated the relationship between cognition and the LC–Hippocampus functional connectivity. These results shed light on some of the earliest brain–behaviour alterations due to risk of future AD in cognitively healthy individuals cross-sectionally. We found a significant longitudinal effect of age on the LC– Hippocampus FC, with no longitudinal effects of risk factors on cognition, the LC– Hippocampus functional connectivity, or their relationship. The lack of prominent longitudinal effects of risk factors is likely due to the narrow follow-up time window (2 years) in this relatively young midlife cohort, an estimated 23 years from dementia onset (Dounavi et al., 2022).

Higher CAIDE scores were significantly associated with worse cognition in verbal, visuospatial functions and short-term (conjunctive) memory, both at baseline and at follow-up. Previous studies from the same cohort have reported significant negative associations of CAIDE scores with visuospatial functions and navigational abilities (Ritchie et al., 2018; Ritchie et al., 2017). We did not observe a longitudinal effect, similarly to other studies of this cohort (Dounavi et al., 2021; Low et al., 2021), possibly due to the relatively young age and the short follow-up window (Ritchie et al., 2018; Ritchie et al., 2017). Post-menopausal women only showed worse cognition with increasing CAIDE scores. The menopausal transition is a middle-age phenomenon with knock-on effects on endocrinal (Weber et al., 2014), cardiovascular (Yanes & Reckelhoff, 2011), and metabolic (Anagnostis et al., 2019; Rahman et al., 2020) systems that significantly impact brain health (Mosconi et al., 2018; Udeh-Momoh & Watermeyer, 2021). The rapid decline in estrogen, a key hormonal change during menopausal transition, is considered to increase vulnerability to AD in females (Scheyer et al., 2018; Udeh-Momoh & Watermeyer, 2021). Post-menopause has been associated with increased levels of Aβ, elevated tau burden, reduced white matter integrity, and glucose hypometabolism (Buckley et al., 2022; Mosconi et al., 2017; Rahman et al., 2020). Our results suggest that post-menopausal status exacerbates the effect of cardiovascular AD risk on cognition, rendering females particularly vulnerable to cognitive decline, and conversely, that addressing cardiovascular risk early may protect cognition in post-menopausal females.

We found that the LC–Hippocampus functional connectivity decreased significantly over two years. The previously reported effect of age on the LC–Hippocampus functional connectivity is inconsistent, possibly due to different age ranges used in different studies. In healthy young and early midlife adults (age range: 18–49), LC functional connectivity to the parahippocampus was negatively associated with age (Zhang et al., 2016). However, another study of a lifespan adult cohort (age range: 19-74) found no association of age with the LC– Hippocampus/parahippocampus functional connectivity (Jacobs et al., 2018). In a broader age-range lifespan cohort (age range: 8–83), Song et al. (2021) found a positive quadratic age effect (u-shape) on LC–parahippocampal functional connectivity, suggesting an increased interaction between these two regions in children and older adults. Therefore, our finding advances the literature, by showing that the LC–Hippocampus functional connectivity decreases with age in midlife.

Another key question here was whether risk factors impact the LC–Hippocampus functional connectivity and its role in cognition. We found that the CAIDE score significantly moderated this brain-behaviour relationship. Stronger LC–Hippocampus functional connectivity was associated with better cognition for individuals with low CAIDE score, and with worse cognition in those with high scores. The presence of this effect in the follow-up, but not the baseline dataset is likely due to the older age of the cohort at follow-up. Previous studies in older adults found that stronger LC–Hippocampus functional connectivity was significantly associated with better cognition (Jacobs et al., 2015). Our finding advances understanding by suggesting that the connectivity between the LC and the hippocampus supports cognition in midlife individuals who have low dementia risk scores.

Furthermore, our result suggests that, in the presence of high dementia risk scores, the LC– hippocampus functional connectivity is negatively associated with cognition. Previous studies have shown that a high CAIDE score, of more than 12, confers a probability of 16.4% for future dementia, with scores above 8 associated with a probability of more than 4% (Kivipelto et al., 2006). In a study of middle-aged individuals with a mean age of 46, participants with a score greater than 8 had a 29% 40-year risk for dementia (Exalto et al., 2014). These findings suggest that a proportion of our study’s participants, who have a CAIDE score greater than 8, may develop dementia. Assuming that AD pathology is underway in this proportion of individuals with high dementia risk scores, our result lends indirect support to the hypothesis that LC hyperactivity and associated hyper-connectivity initiates the spread of pathological tau to MTL (Weinshenker, 2018), and particularly the hippocampus, in the early stages of AD. The ensuing neurodegeneration may explain why hyper-connectivity is associated with worse performance in individuals with high dementia risk scores. However, the neural mechanisms of AD neuropathology genesis remain contested. Given the lack of tau biomarker status for this cohort, future studies that integrate tau and functional connectivity information are required to further test the proposal that tau spread from the LC to the hippocampus via functional connectivity pathways. Furthermore, future studies from the continuing longitudinal follow-up of this cohort will validate the role and nature of these changes in individuals who display AD neuropathology as they get older.

We did not observe any significant effects of risk factors for AD on the LC–Hippocampus connectivity. Previous studies on MCI and AD patients have shown disrupted/lower LC– Hippocampus functional connectivity (MCI: Jacobs et al., 2015; age: 65.1 ± 4.5 years) (Liebe et al., 2022; age: 73.3 ± 7.5 years; AD patients: Zhao et al., 2017), compared to age-matched healthy controls. One previous study investigating cognitively healthy middle-aged adults with familial risk for late-onset dementia found no effect of AD risk on the LC–Hippocampus connectivity (Del Cerro et al., 2020; age: 50.4 ± 8.3 years). Consistent with this previous study, our null result suggests that AD risk factors are not associated with the LC– Hippocampus connectivity in middle-aged individuals who are cognitively healthy.

We also observed a trend effect association of APOE ε4 allele with better cognition in episodic and relational memory, independently of sex, age and years of education. Previous research has suggested that the APOE ε4 allele, although being associated with poorer health outcomes in old age, may have a positive effect earlier in life. For example, it has been associated with better visual perception (Ritchie et al., 2017), short-term memory (Zokaei et al., 2020), higher intelligence quotient and a more economic use of memory-related neural resources in young healthy humans (Mondadori et al., 2007). These results may also be interpreted based on the antagonistic pleiotropy hypothesis of aging (William 1957), which proposes that deleterious genes, such as the APOE ε4 gene allele, have survived through evolution because they might confer an advantage early in life, when humans are reproductively fit. These results suggest an impact of genetic risk for late-life sporadic AD in cognitively healthy midlife individuals on episodic memory, the hallmark cognitive dysfunction in early clinical dementia and mild cognitive impairment (Irish et al., 2011). Thus, they highlight the importance of studying the origins of Alzheimer’s disease in this age group, over 20 years before typical symptom onset. Given the weak effect however, further replication in larger samples is warranted to validate these findings.

In summary, the PREVENT-Dementia study is a longitudinal multi-site study targeted at identifying early biomarkers for Alzheimer’s disease. In this study, we provide early proof that the recruited cohort, with a mean age of 52/54 years at baseline/follow-up, showed a disrupted role of LC–Hippocampus connectivity in cognition with an increasing CAIDE score. APOE ε4 genotype or FHD did not moderate this brain–behaviour relationship. In other words, when the genetic risk alone was considered, no alterations of brain–behaviour relationships were found. It was only when a risk score (CAIDE) incorporating genetic risk in combination with lifestyle factors, sex and age was considered, that such alterations were unravelled. Taken together, these results provide strong evidence that brain–behaviour alterations in individuals with higher CAIDE scores may be driven by lifestyle risk factors included in this dementia risk score (i.e., blood pressure, cholesterol, physical activity, body mass index, years of education).

Hence, these findings highlight the importance of considering modifiable risk factors when stratifying risk populations, or potentially designing randomized control trials. Our previous work in this cohort has demonstrated that modifiable lifestyle factors affect cognition, particularly in individuals at high risk for late-life AD (Heneghan et al., 2023) and lends further support to this idea. In fact, a randomized multi-domain control trial in the FINGER population (mean age 70 years old) over two years demonstrated that the applied lifestyle and vascular interventions had an impact on cognition (Stephen et al., 2019). Hence, further investigation of pathological alterations in relation to modifiable risk factors in midlife is warranted to unveil the sequalae of brain and behaviour alterations leading to dementia.

### 4.1 Strengths and Limitations

Strengths of this study are its large well-characterized middle-aged cohort. State-of-the-art analysis methods and a thorough quality control protocol were applied to the acquired data. Limitations include the short follow-up window of the study and the absence of further well-established preclinical biomarkers, such as amyloid and tau status. The interpretability of the effect of risk on individual cognitive abilities requires individuation in future studies. Standard MR functional sequences provide limited resolution of the diminutive LC volume, and thus, higher resolution imaging that specifically targets the LC region is required to further validate these findings in future studies. Like most other studies involving mid-life and older people in the UK and Ireland, the study population is mainly (95%) of white Caucasian ethnicity, not dissimilar to the historic ethnic mix of older people in the five study sites in this region. The inclusion of other ethnicities and representation from different world regions in future studies will ensure true diversity and findings that can be generalized to different populations across the globe.

## Data Availability

Data are available on reasonable request.

## Acknowledgement

F.D. was funded by the Provost PhD Award Scheme from Trinity College Dublin, to L.N. L.N. was also funded by a L’Oréal-UNESCO for Women In Science International Rising Talent Award, the Welcome Trust Institutional Strategic Support grant, and the Global Brain Health Institute Project Grant.

The PREVENT-Dementia study is supported by the UK Alzheimer’s Society (grant numbers 178 and 264), the US Alzheimer’s Association (grant number TriBEKa-17-519007) and philanthropic donations.

We thank all PREVENT-Dementia participants for their enthusiastic participation in this study. We also thank the Research Delivery service at West London NHS Trust and the Wolfson Clinical Imaging Facility at Imperial College London for their support in running the study.

