## Supporting Information File for "Cardiovascular risk of dementia is associated with brain–behaviour changes in cognitively healthy, middle-aged individuals"

**Title**

**Affiliations**

School of Psychology

Trinity College Institute of Neuroscience

Global Brain Health Institute

Trinity College Dublin

Dublin, Ireland

**Materials and Methods**

The eleven cognitive summary variables from the COGNITO battery were:

1. *Working memory*. The ‘double task’ requires the subject to simultaneously locate specific shapes (visual stimuli) and count presented sounds (auditory stimuli). The outcome variable is the total number of correct responses for shape recognition.
2. *Working memory*. This task consists of two parts - the ‘double task’ described above and a visual task. The visual task requires the subject to locate specific shapes (visual stimuli) without the presentation of auditory stimuli. The outcome variable is the mean time difference in milliseconds between the 'double task' and the the visual task.
3. *Narrative recall*. The task requires the subject to recall a series of elements which have a logical sequence (a short story). The outcome variable is the total number of correct elements on immediate recall of a story with a temporal progression requiring attention to macrostructure.
4. *Description recall*. The task requires the subject to recall a series of elements that have a visual sequence (e.g., describing the layout of a house).The outcome variable is the total number of correct elements recalled of a description without thematic progression requiring attention to microstructure and recall of spatial location. The narrative and description recall tasks are similar in terms of word frequency in the language and syntactic structure.
5. *Implicit memory*. The task requires the subject to recognise as soon as possible a name that is constructed progressively on the screen in 15 steps. The outcome variable is the difference in the number of steps required for recognition between names never seen and names previously learned.
6. *Name-face association*. Participants are presented with a set of faces and names and the goal is to remember and correctly associate each face with its corresponding name. During the learning phase, participants are required to learn the associations between the faces and names, which they are then required to recall after a delay. During the recall phase participants are shown a series of faces, some of which have been shown previously and associated with names. Participants decide whether the face has appeared before. If participants say they have seen the face before, they are asked for the name. The outcome variable is the number of names correctly recalled.
7. *Form perception*. The task requires the subject to discriminate form and line orientation by matching a sample complex figure to one of six presented figures. The outcome variable is the number of correct responses.
8. *Form perception speed*. The task is the same as above. The outcome variable is the mean time of correct responses in milliseconds.
9. *Phoneme comprehension*. The task requires the subject to select one of six presented objects that illustrates a word. The six objects include shape, phonetic and semantic distractors. The outcome is the number of correct responses.
10. *Phoneme comprehension speed*. The task is the same as above. The outcome variable is the mean time of correct responses in milliseconds.
11. *Verbal fluency*. The task requires the subject to name as many objects as they can think of, given a semantic and a phonemic cue. The outcome variable is the total sum of the number of words generated in 60s using both a semantic (vegetables) and a phonemic (letter P) cue.

The two summary variables from the Visual Short-term Memory Binding task were:

12. *Visual short-term memory binding test (shape only condition)*. The task requires the subject to identify whether the shape of the test stimuli (three random 6-sided polygons) matches the studied stimuli. The outcome variable is the percentage of correctly recognised shapes after a short retention period.
13. *Visual short-term memory binding test (shape-color binding condition)*. The task requires the subject to identify whether both the shape and colour of the test stimuli match the studied stimuli. The outcome variable is the percentage of correct recognition of combinations of shape and colour of the test stimuli after a short retention period.

**Behavioural data reduction**

Sharp breaks in the “scree" plot of the successive eigenvalues suggest the appropriate number of components to extract. We also conducted a parallel analysis that compares the scree of components of the actual data with that of a random data matrix of the same size as the original (Horn, 1965). The detailed steps are: 1) We simulated a random normal data matrix of the same number of original cognitive variables (n=13) and the same number of participants (n=208 at baseline and n=166 at follow-up). 2) We extracted eigenvalues from the simulated data matrix. We repeated these two steps 500 times to create a set of 500 parallel eigenvalues. 3) We took the mean and 95th percentile of all eigenvalues generated by principal components analysis of random data sets. The results were a vector of mean (and 95th percentile) eigenvalues equal in size to the original number of cognitive variables (n=13). 4) We compared the eigenvalues of the actual data to that of parallel random data sets. Specifically, we plotted eigenvalues from the actual and random data sets and kept only those components whose eigenvalues are greater than 95th percentile of eigenvalues from the random data sets. Figure 2a in the manuscript shows the scree plots of eigenvalues based on the actual data matrix (in blue line) and simulated data matrices at baseline (left panel) and follow-up (right panel) The mean eigenvalues are in dashed grey line, and the 95th percentile of eigenvalues are in red line. The shaded areas indicate that the eigenvalues of components based on the actual data were larger than the mean and 95th percentile of eigenvalues from the random data sets. Figure 2b shows the proportion of variance that each component explained, and the cumulative variance of the three components.

**Framewise displacement calculation**

Differentiating head realignment parameters across frames yields a six dimensional timeseries that represents instantaneous head motion, which can then be summarised as a scalar quantity, framewise displacement (FD), using the formula (Equation (1)). Specifically, this measure was calculated as the sum of the absolute values of the derivatives of the 6 realignment parameters (Power et al., 2012). Rotational displacements were converted from degrees to millimetres by calculating the displacement on the surface of a sphere with a radius of 50 mm, which is approximately the mean distance from the cerebral cortex to the centre of the head.

$$\boldsymbol{FD}_{\boldsymbol{i}}\boldsymbol{=}\left| \boldsymbol{\Delta}\boldsymbol{d}_{\boldsymbol{ix}} \right|\boldsymbol{+}\left| \boldsymbol{\Delta}\boldsymbol{d}_{\boldsymbol{iy}} \right|\boldsymbol{+}\left| \boldsymbol{\Delta}\boldsymbol{d}_{\boldsymbol{iz}} \right|\boldsymbol{+}\left| \boldsymbol{\Delta}\boldsymbol{\alpha}_{\boldsymbol{i}} \right|\boldsymbol{+}\left| \boldsymbol{\Delta}\boldsymbol{\beta}_{\boldsymbol{i}} \right|\boldsymbol{+}\left| \boldsymbol{\Delta}\boldsymbol{\gamma}_{\boldsymbol{i}} \right|\boldsymbol{(1)}$$

Where $\Delta\boldsymbol{d}_{\boldsymbol{ix}}\boldsymbol{=}\boldsymbol{d}_{\left( \boldsymbol{i-1} \right)\boldsymbol{x}}\boldsymbol{-}\boldsymbol{d}_{\boldsymbol{ix}}$  and similarly for the other rigid body parameters $\left[ \boldsymbol{d}_{\boldsymbol{iy}}\boldsymbol{,}\boldsymbol{d}_{\boldsymbol{iz}}\boldsymbol{,}\boldsymbol{\alpha}_{\boldsymbol{i}}\boldsymbol{,}\boldsymbol{\beta}_{\boldsymbol{i}}\boldsymbol{,}\boldsymbol{\gamma}_{\boldsymbol{i}} \right]$**.**

**Results**

**Cognitive components**

The components can be interpreted by mapping out the cognitive functions that the highest loading measures tapped into (Figure 3a). For the first component (C1), given that the highest loading measures were assessing response accuracy, and were positively loaded on C1, higher value means better performance. The cognitive functions that the highest three loading measures tapped into were verbal (narrative recall), spatial (description recall) and relational memory (name-face association). Accordingly, C1 was labelled as ‘episodic and relational memory’. For the second component (C2), the highest loading measures were assessing both response accuracy and latency. As accuracy measures were positively, and latency measures were negatively loaded on C2, higher value means better performance. Based on the cognitive functions that the highest loading measures tapped into, C2 was labelled as ‘working and short-term (single-feature) memory’. For the third component (C3), the highest loading measures were also assessing both response accuracy and latency. By contrast to C2, accuracy measure was negatively, and latency measures were positively loaded on C3. Therefore, higher value means poorer performance. For clarity of graphical display and comparison to the other two, we reversed the signs of the values to make larger values represent better performance. Based on cognitive functions that the highest loading measures tapped into, C3 was labelled as ‘verbal, visuospatial functions, and short-term (conjunctive) memory’.

**Associations of the mean framewise displacement with risk factors and LC–Hippocampus functional connectivity**

After carefully censoring the motion-contaminated data, we found no significant difference in mean FD between APOE ε4 carriers and non-carriers at either baseline (t = 1.05, p = 0.30) or follow-up (t = 1.40, p = 0.17), nor between FHD+ and FHD- at either time point (baseline: t = -0.32, p = 0.75; follow-up: t = 0.51, p = 0.61). There was also no significant correlation between mean FD and CAIDE at either baseline (r = 0.07, p = 0.44) or follow-up (r = 0.08, p = 0.43). Furthermore, mean FD was not significantly associated with LC–Hippocampus functional connectivity at either time point (baseline: r = 0.01, p = 0.91; follow-up: r = 0.05, p = 0.60). These results suggest that head movement did not significantly influence our results.

**Validation analyses with APOE** ε**2**ε**4 carriers excluded**

The prevalence of APOE ε2ε4 in our cohort is 0.02% (SFigure 2). To test whether our main findings were confounded by the inclusion of APOE ε2ε4 carriers, we also performed analyses excluding APOE ε2ε4 carriers. We found a significant positive association of the APOE ε4 allele with episodic and relational memory at baseline [β (SE) = 0.27 (0.14), p = 0.05] and at follow-up [β (SE) = 0.30 (0.15), p = 0.05], independent of sex, age and years of education. APOE ε4 carriers performed significantly better than non-carriers. No significant associations of the APOE ε4 allele with performance in the other two cognitive domains were observed. These results were consistent with our main findings including APOE ε2ε4 carriers (SFigure 6) (Table 2).

In addition, Spearman correlation analyses showed a significant negative association of CAIDE (including APOE genotype) with verbal, visuospatial functions and short-term (conjunctive) memory at baseline (rho = -0.16, p = 0.02) and at follow-up (rho = -0.19, p = 0.01). Higher CAIDE was significantly associated with poorer cognitive performance. No significant associations of CAIDE with the other two cognitive domains were observed. These results were also consistent with our main findings when we used CAIDE scores that included APOE ε2ε4 allele (Figure 4).

Additionally, there was no significant association between any of the risk factors and the LC–Hippocampus functional connectivity, either cross-sectionally or longitudinally, which is also consistent with our main findings, with APOE ε2ε4 carriers included.

Lastly, CAIDE (including APOE genotype) showed a significant moderation effect on the relationship between LC–Hippocampus functional connectivity and cognition comprising verbal, visuospatial functions and short-term (conjunctive) memory at follow-up [β (SE) = -0.56 (0.24), p = 0.02], consistent with our main findings when we used CAIDE scores including the APOE ε2ε4 allele (Figure 6) (Table 5).

Taken together, these results suggest that the inclusion of APOE ε2ε4 carriers did not affect our main results.

**Demographic characteristics of the cohort stratified by sex**

Demographic characteristics of the cohort at baseline and follow-up, stratified by sex, are shown in STable 3. There were no significant differences in age, years of education, FHD, or APOE ε4 allele genotype between males and females. By design, CAIDE scores were significantly higher in males than in females at baseline (*p* = 0.0001) and at follow-up (*p* = 0.0001), as male is scored higher in the calculation of CAIDE scores (STable 3).

**Impact of outliers**

To assess whether the main results were driven by outliers, we first displayed scatterplots of the cognitive variables, LC–Hippocampus functional connectivity and hippocampus volume to show the distribution of the data at baseline (SFigure 4) and at follow-up (SFigure 5), and then used both Tukey's fences and z-scores to examine outliers, providing a comprehensive analysis of data points that deviated significantly from the central tendency. Tukey's fences involve the use of the interquartile range (IQR) to set upper and lower limits beyond which data points are considered potential outliers. Z-scores, on the other hand, standardise data points based on the mean and standard deviation, identifying observations that are significantly outside the mean. We set the threshold for Tukey's fences at 1.5 times the IQR, and for z-scores we considered data points beyond ±3 standard deviations. These techniques allow for a systematic and nuanced examination of outliers, contributing to a more robust analysis of the data distribution.

Lastly, we re-examined the main results with the identified outliers excluded. In SFigure 6, we identified two outliers in APOE ε4 carriers at follow-up, but no outliers at baseline. This suggests that the significant effect of the APOE ε4 allele on episodic and relational memory at baseline was not driven by outliers. After excluding the outliers at follow-up, the trend effect of APOE ε4 genotype disappeared. In Figure 4 there are two outliers at follow-up (Figure 4b) but none at baseline (Figure 4a). After excluding these outliers, the negative association between CAIDE and verbal, visuospatial functions, and short-term (conjunctive) memory remained significant at follow-up (rho = -0.21, p = 0.006). In the updated Figure 5, we detected an outlier at baseline and then re-examined the results after excluding this outlier. The longitudinal change in LC–Hippocampus functional connectivity over 2 years remained significant [β (SE) = -0.06 (0.03), p = 0.05]. In Figure 6 we found three outliers at baseline and one at follow-up. The interaction between CAIDE and LC–Hippocampus functional connectivity on cognition at follow-up remained significant after excluding the outlier [β (SE) = -0.43 (0.21), p = 0.04]. In summary, these analyses show that the main results are robust and unaffected by the presence of extreme values.

**Figures and Legends**

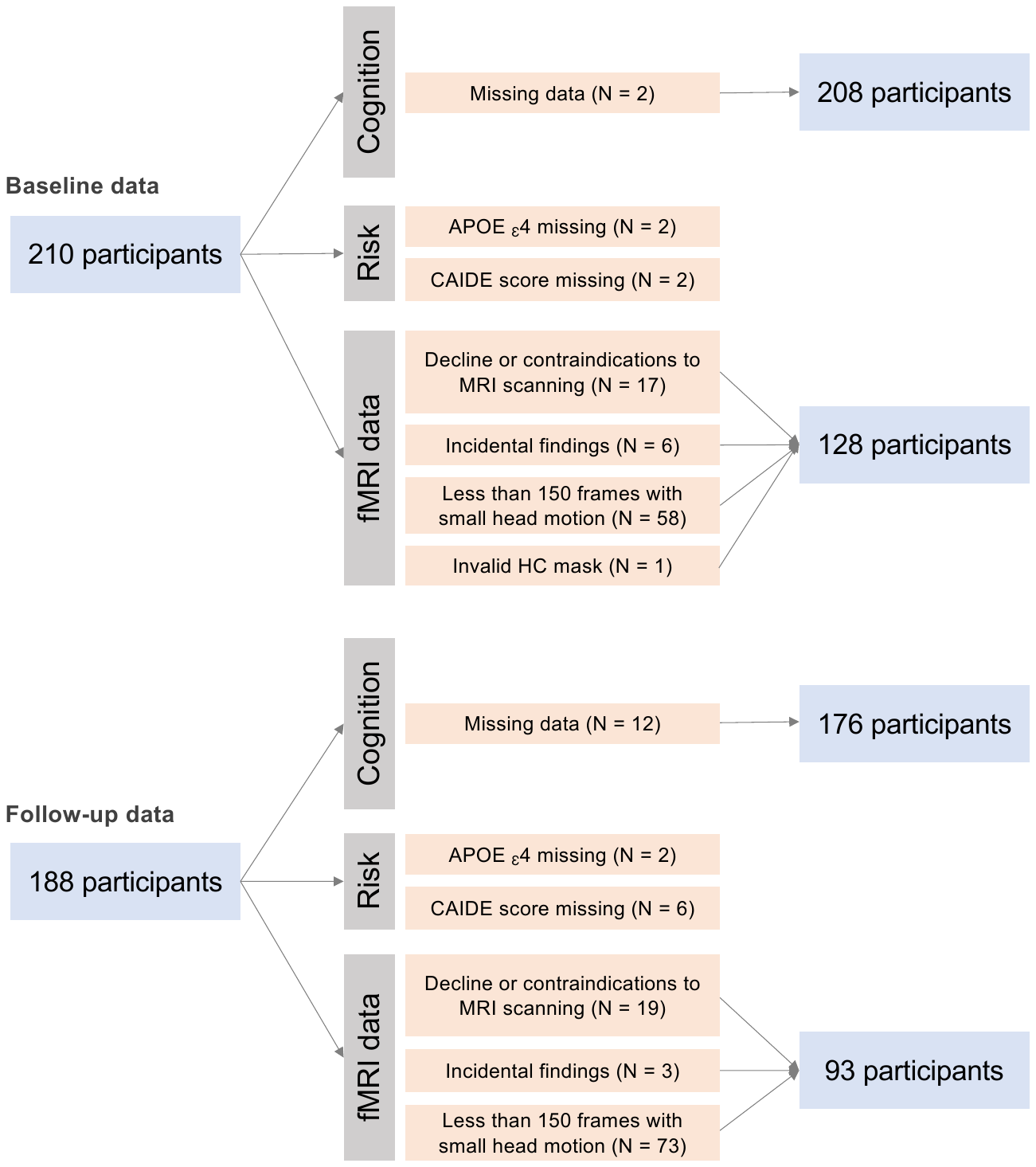

SFigure 1. Participant inclusions for different analyses.

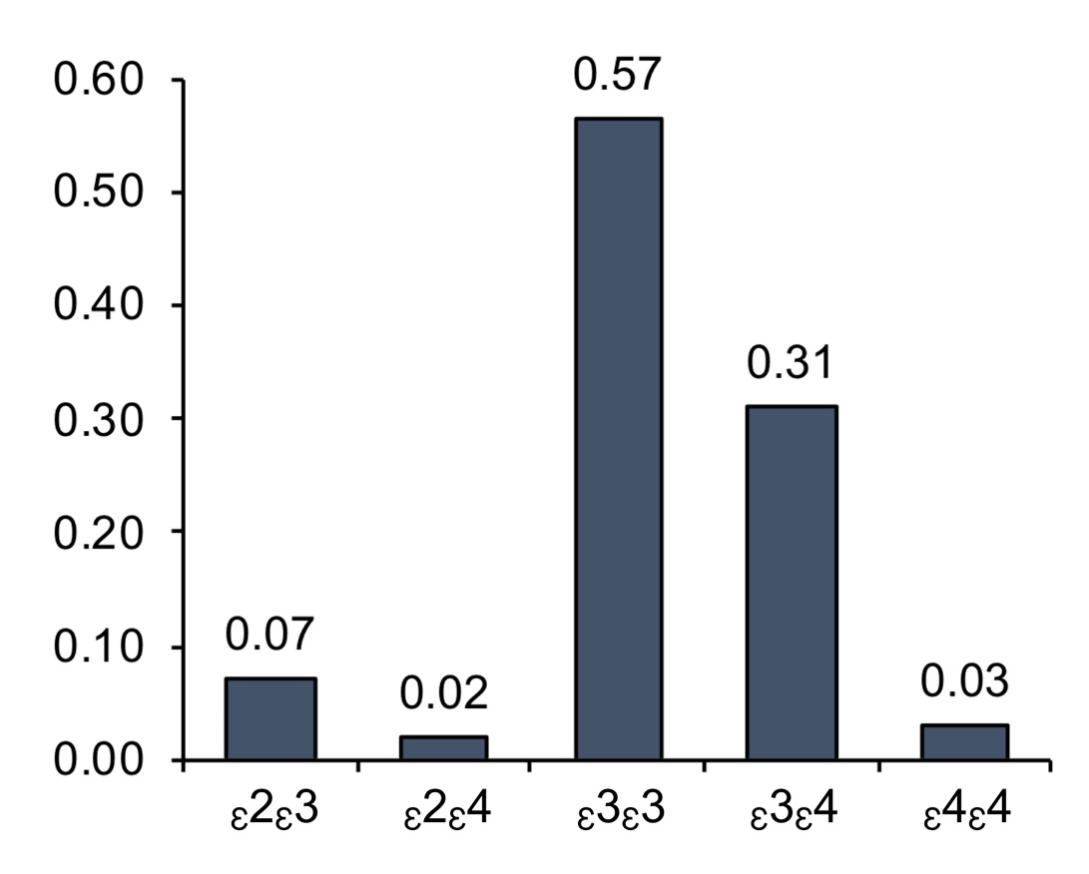

SFigure 2. The prevalence of APOE genotype.

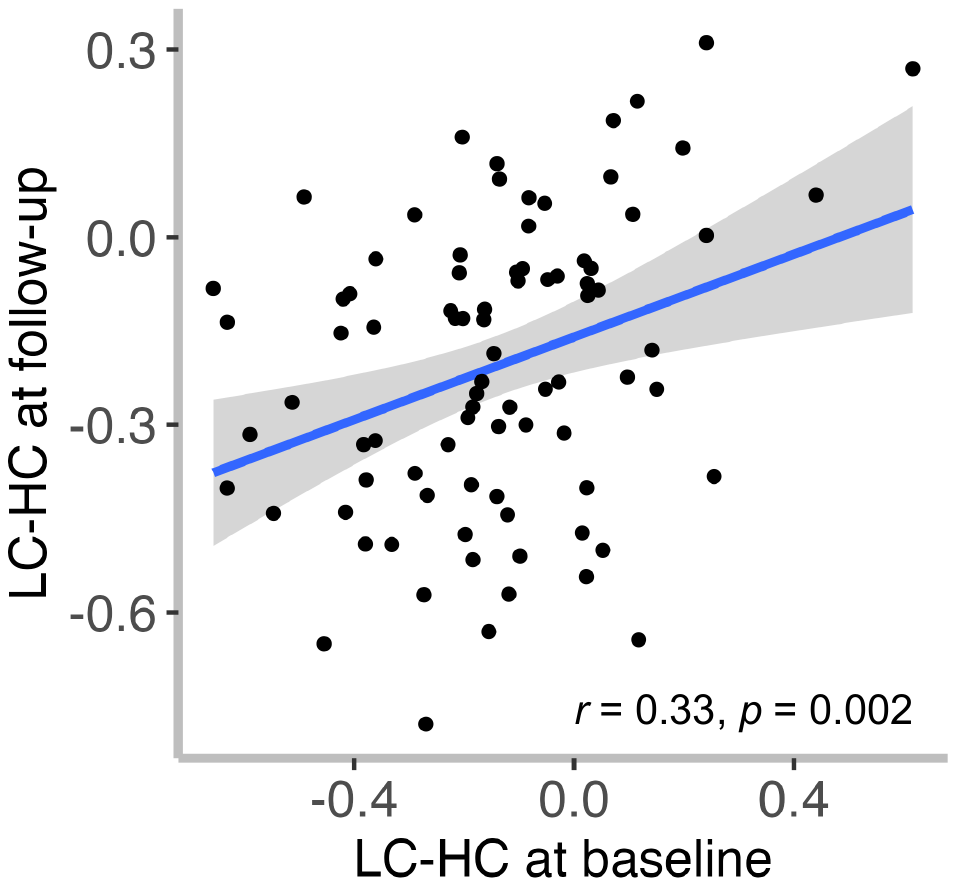

SFigure 3. There was a significant positive correlation of the LC–Hippocampus functional connectivity between the two study time points (r = 0.33, p = 0.002).

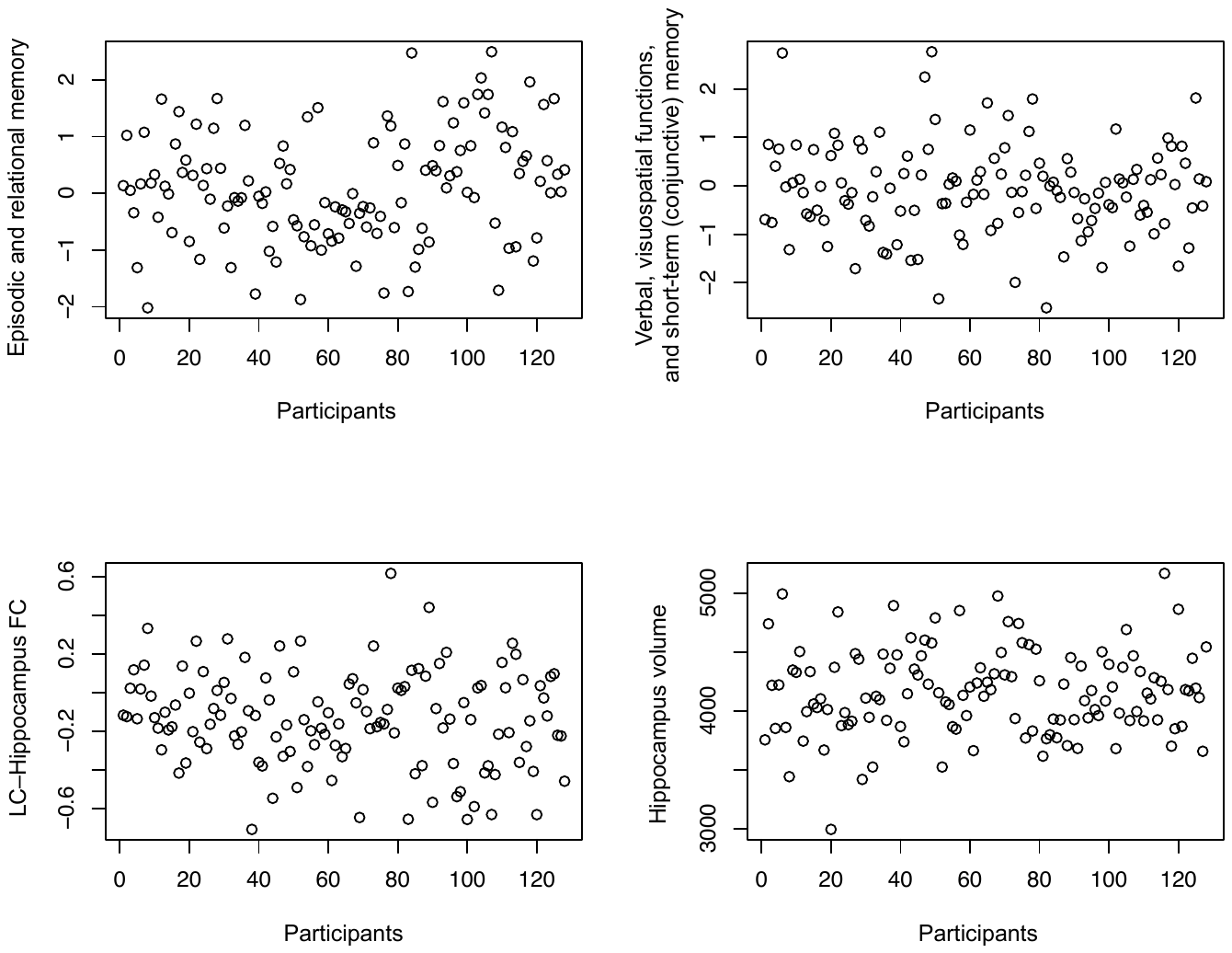

SFigure 4. Scatterplots of the cognition, LC–Hippocampus functional connectivity (FC) and the hippocampus volume at baseline.

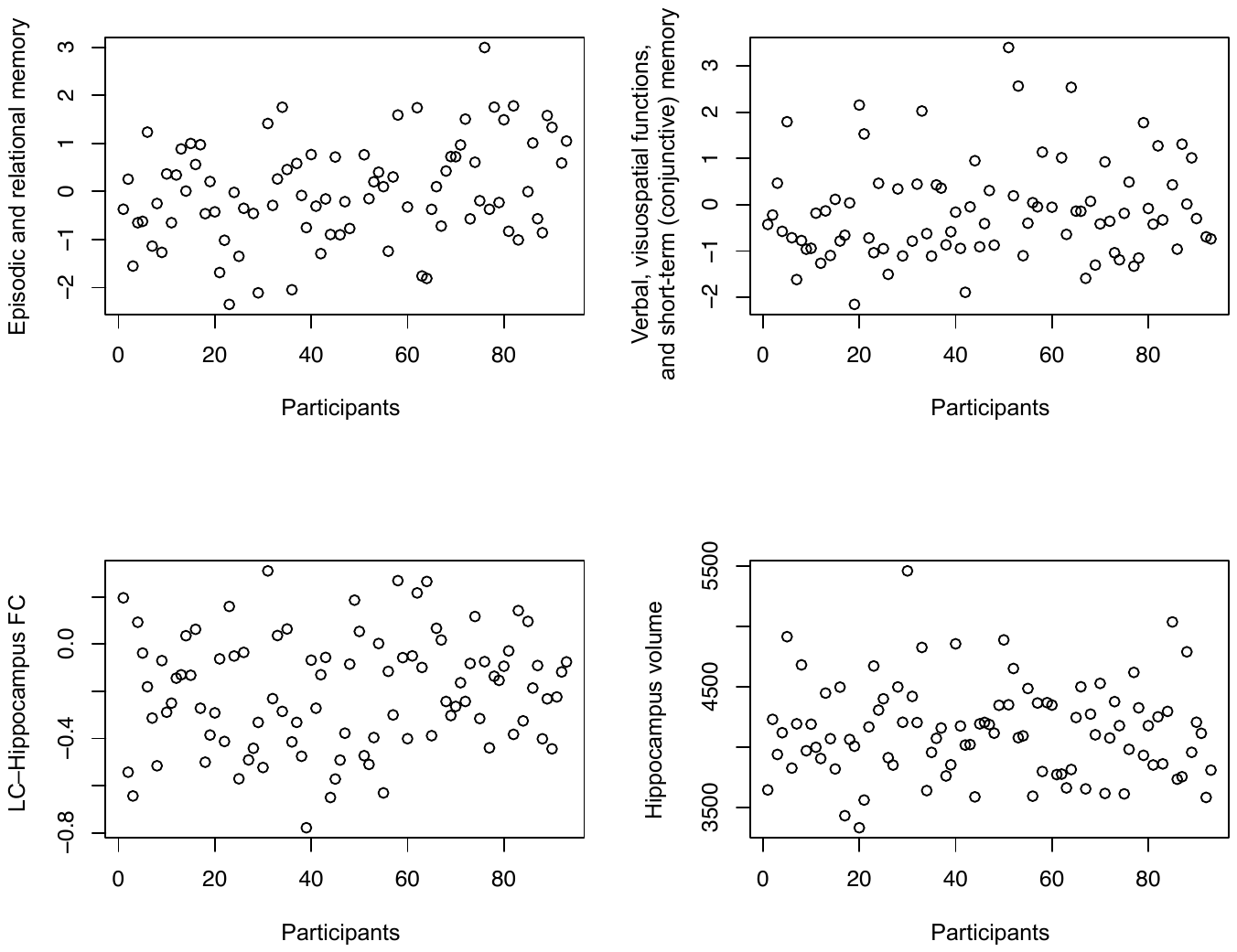

SFigure 5. Scatterplots of the cognition, LC–Hippocampus functional connectivity (FC) and the hippocampus volume at follow-up.

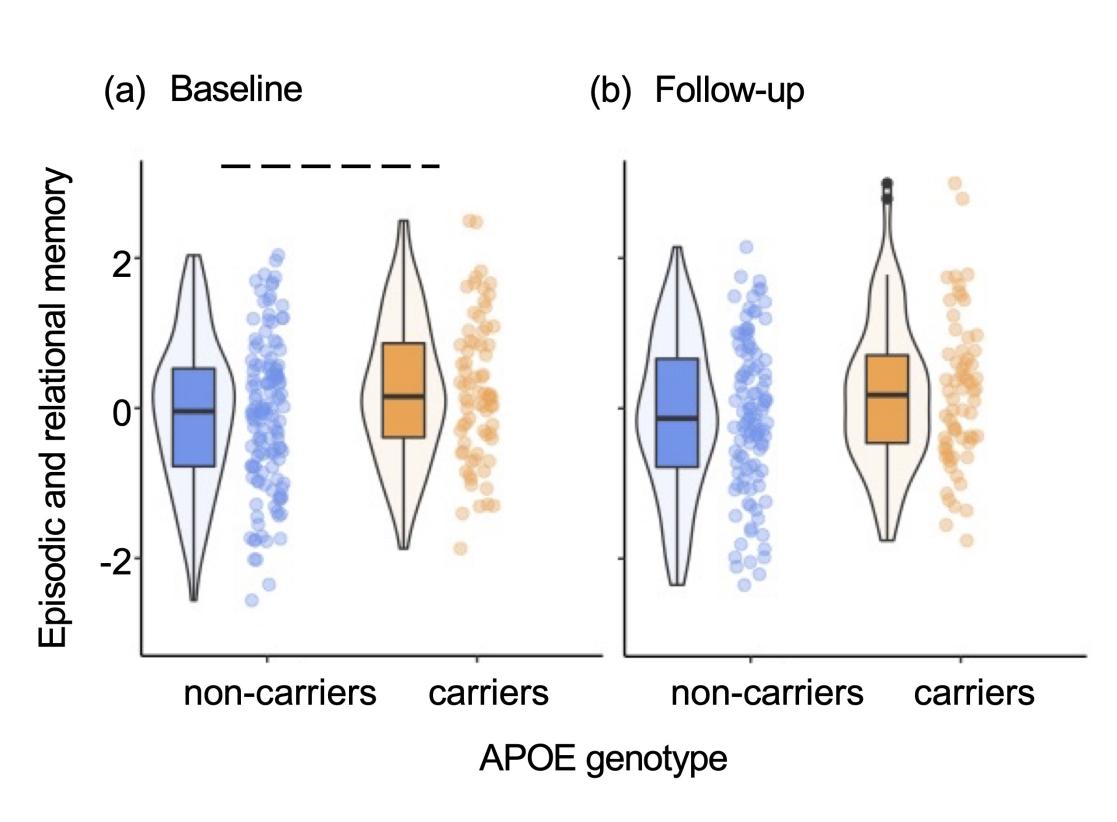

SFigure 6. The effect of APOE genotype on episodic and relational memory (a) at baseline and (b) follow-up. We observed a trend effect (dashed line) at baseline when corrected for multiple comparisons.

STable 1. Complete list of risk factors and tests obtained at baseline and follow-up

| Baseline | Follow-up |
| --- | --- |
| **Risk factors** | **Risk factors** |
| *Family history* | *Family history* |
| *APOE* ε*4* (Missing = 2) | *APOE* ε*4 (*Missing = 2) |
| *CAIDE (*Missing = 2) | *CAIDE (*Missing = 6) |
| Age | Age |
| Sex | Sex |
| Yeas of education | Yeas of education |
| Systolic blood pressure | Systolic blood pressure |
| BMI | BMI (Missing = 1) |
| Cholesterol | Cholesterol (Missing = 2) |
| Physical activity | Physical activity (Missing = 1) |
| APOE ε4 (Missing = 2) | APOE ε4 (Missing = 2) |
| **Neuropsychological assessments** | **Neuropsychological assessments** |
| *COGNITIO* | *COGNITIO* (Missing = 4) |
| Working memory | Working memory |
| Narrative recall | Narrative recall |
| Description recall | Description recall |
| Implicit memory | Implicit memory |
| Name-face association | Name-face association |
| Form perception | Form perception |
| Phoneme comprehension | Phoneme comprehension |
| Verbal fluency | Verbal fluency |
| *VSTMBT* (Missing = 2) | *VSTMBT* (Missing = 8) |
| **Neuroimaging data**  *fMRI* (Missing = 17) | **Neuroimaging data**  *fMRI* (Missing = 19) |

| STable 2. Description of cognitive tasks and measures | | | | |
| --- | --- | --- | --- | --- |
| Cognitive Tasks | Measures | Task Description | Descriptive Statistics (N=210, baseline) | Descriptive Statistics (N=188, follow-up) |
| Working memory | Total number of correct answers | Dual task: The subject must locate the targeted shapes and count the sounds. | Mean = 9.75 SD = 0.54 Range = 3.00 Missingness = 0 | Mean = 9.85 SD = 0.51 Range = 3.00 Missingness = 4 |
|  | Mean time (ms) difference in milliseconds between the dual task and a simple form recognition task | Dual task: The subject must locate the targeted shapes and count the sounds.  Simple task: Subject must locate the targeted shapes only. | Mean = 223.09 SD = 3175.07 Range = 18238.00 Missingness = 0 | Mean = -55.47 SD = 2919.28 Range = 17750.00 Missingness = 4 |
| Narrative recall | Total number of correct answers | The subject must recall a series of elements which have a logical sequence (a short story). | Mean = 13.25 SD = 4.73 Range = 23.00 Missingness = 0 | Mean = 15.03 SD = 4.30 Range = 23.00 Missingness = 4 |
| Description recall | Total number of correct answers | The subject must recall a series of elements which have a visual sequence (a short description). | Mean = 12.63 SD = 4.34 Range = 20.00 Missingness = 0 | Mean = 13.39 SD = 4.73 Range = 25.00 Missingness = 4 |
| Implicit memory | Difference the number of names never seen and the number of names already learned | The subject must recognize as soon as possible a name which is constructed progressively on the screen. | Mean = 1.05 SD = 0.83 Range = 8.20 Missingness = 0 | Mean = 1.11 SD = 0.69 Range = 5.00 Missingness = 4 |
| Name-face association | The number of correctly recognized names | The subject must decide whether a face on the screen appeared before and if yes, what the person's name is. | Mean = 5.37 SD = 2.21 Range = 9.00 Missingness = 0 | Mean = 5.86 SD = 2.02 Range = 9.00 Missingness = 4 |

| STable 2. Description of cognitive tasks and measures (continued) | | | | |
| --- | --- | --- | --- | --- |
| Form matching | Total number of correct answers | The subject must discriminate form and line orientation by matching a sample complex figure to one of six figures. Distractor figures are designed to detect visuospatial field neglect and difficulties with line orientation. | Mean = 6.45 SD = 1.12 Range = 7.00 Missingness = 0 | Mean = 6.43 SD = 1.01 Range = 4.00 Missingness = 4 |
|  | Mean time (ms) for correct answers |  | Mean = 5841.64 SD = 1425.88 Range = 7596.00 Missingness = 0 | Mean = 6071.80 SD = 1567.05 Range = 8116.00 Missingness = 4 |
| Phoneme comprehension | Total number of correct answers | The subject must choose an object illustrating a presented word among 6 objects which include shape, phonetic and semantic distractors. | Mean = 8.62 SD = 0.57 Range = 3.00 Missingness = 0 | Mean = 8.63 SD = 0.53 Range = 2.00 Missingness = 4 |
|  | Mean time (ms) for correct answers |  | Mean = 1585.71 SD = 304.24 Range = 1654.00 Missingness = 0 | Mean = 1508.96 SD = 272.53 Range = 1724.00 Missingness = 4 |
| Verbal fluency | Total number of correct answers | The subject must name all the words they can think of within one minute based on semantic and phonetic cues. | Mean = 28.39 SD = 6.58 Range = 32.00 Missingness = 0 | Mean = 29.59 SD = 7.12 Range = 43.00 Missingness = 4 |
| VSTMBT-shape only | Total number of correct answers | The subject must recall stimuli that were shapes after a short period of retention. | Mean = 0.85 SD = 0.14 Range = 0.88 Missingness = 2 | Mean = 0.86 SD = 0.17 Range = 2.31 Missingness = 10 |
| VSTMBT-shape colour binding | Total number of correct answers | The subject must recall stimuli that were combinations of shapes and colours after a short period of retention. | Mean = 0.52 SD = 0.20 Range = 1.19 Missingness = 2 | Mean = 0.54 SD = 0.24 Range = 2.19 Missingness = 11 |
| Key: VSTMBT, visual short-term memory binding test; SD, standard deviation | | | | |

STable 3. Demographic characteristic of the cohort at baseline and follow-up based on biological sex

|  | Baseline | | | Follow-up | | |
| --- | --- | --- | --- | --- | --- | --- |
|  | Female | Male | *p* | Female | Male | *p* |
|  | (n=148) | (n=62) | (Mann-Whitney U) | (n=133) | (n=55) | (Mann-Whitney U) |
| Age (y) | 52.0 ± 8.3 | 53.0 ± 8.0 | 0.17 | 55.0 ± 7.0 | 55.0 ± 8.5 | 0.31 |
| Years of Education | 16.0 ± 5.0 | 16.0 ± 3.0 | 0.94 | 17.0 ± 5.0 | 16.0 ± 3.5 | 0.75 |
| CAIDE  (incl. APOE status) | 5.0 ± 3.0 | 7.0 ± 3.8 | 0.0001 | 6.0 ± 3.0 | 7.0 ± 3.0 | 0.0001 |
|  | (n=146) | (n=62) |  | (n=127) | (n=55) |  |
|  |  |  | *p* (Chi-Square) |  |  | *p* (Chi-Square) |
| Menopausal status  (% Postmenopausal) | 37.16% | - | *-* | 38.35% | - | *-* |
| FHD (% Positive) | 50.00% | 46.77% | 0.78 | 53.38% | 50.91% | 0.88 |
| APOE ε4 (% Carriers) | 36.30% | 35.48% | 1.00 | 35.88% | 38.18% | 0.90 |
|  | (n=146) | (n=62) |  | (n=131) | (n=55) |  |

Note: median ± interquartile range (IQR) was reported for continuous variables. Abbreviations: FHD-, Negative family history of dementia; FHD+, positive family history of dementia; APOE ε4, Apolipoprotein ε4 genotype; CAIDE, Cardiovascular Risk Factors, Aging and Dementia.
